# Leveraging Large-Scale Biobanks for Therapeutic Target Discovery

**DOI:** 10.1101/2025.02.10.25321487

**Authors:** Brian R Ferolito, Hesam Dashti, Claudia Giambartolomei, Gina M Peloso, Daniel J Golden, Kai Gravel-Pucillo, Danielle Rasooly, Andrea RVR Horimoto, Rachael Matty, Liam Gaziano, Yi Liu, Ines A Smit, Barbara Zdrazil, Yakov Tsepilov, Lauren Costa, Nicole Kosik, Jennifer E Huffman, Gian Gaetano Tartaglia, Giorgio Bini, Gabriele Proietti, Harris Ioannidis, Fiona Hunter, Gibran Hemani, Adam S Butterworth, Emanuele Di Angelantonio, Claudia Langenberg, Maya Ghoussaini, Andrew R Leach, Katherine P Liao, Scott Damrauer, Luis E Selva, Stacey Whitbourne, Philip S Tsao, Jennifer Moser, Tom Gaunt, Tianxi Cai, John C Whittaker, Million Veteran Program, Juan P Casas, Sumitra Muralidhar, J Michael Gaziano, Kelly Cho, Alexandre C Pereira

## Abstract

Large biobanks, including the Million Veteran Program (MVP), the UK Biobank, and FinnGen, provide genetic association results for more than 1,000,000 individuals for hundreds of phenotypes. To select targets for pharmaceutical development, as well as to improve the understanding of existing targets, we harmonized these studies, and performed two-sample Mendelian Randomization (MR) on 2,003 phenotypes using genetic variants associated with gene expression (derived from GTEx and eQTLGen) and plasma protein levels (derived from ARIC, Fenland, and DeCODE) as proxies of target modulation. We found 69,669 gene-trait pairs with evidence (p ≤ 1.6 x 10^-9^) for causal effects. From the selected gene-trait pairs, we observed 6,447 genes with strong causal evidence for at least one of 2,003 investigated traits. As expected, being identified as a gene-trait pair in our approach was significantly associated with higher odds of being an approved drug target and indication. We were able to rediscover 9% of approved drug targets in ChEMBL 34. Moreover, identified gene-traits were significantly associated with higher odds of being previously described as a gene-trait pair in OMIM, ClinVar, mouse knock-out data, and rare variant burden studies. To enhance the translational potential of the resource, we developed a predictive ranking model trained using approved drug targets described in ChEMBL 34 as well as several different biological annotations. This model was able to accurately predict the odds of a particular significant MR result being developed into an approved drug and its clinical indication (precision-recall AUC 0.79). We make our results publicly available in <u>CIPHER</u>.

## Introduction

Use of genetic association information in the realm of both common and rare genetic variation to guide drug target identification has been shown to provide significant additional information for drug development programs. In recent years, the availability of large-scale genetic resources such as the Million Veteran Program^1^ (MVP), the UK Biobank^2,3^, and FinnGen^4^ have made it possible to associate millions of genetic variants to thousands of human phenotypes at a scale and pace previously unfathomable^5^. Importantly, these traits include molecular phenotypes such as transcriptomics and proteomics^6,7^. These efforts are having a significant impact on our understanding of the mechanistic determinants of complex human traits, our capacity to predict medical conditions, and our ability to identify drug targets that modulate the causes of these disorders^8,9^.

Despite these advances, there are substantial hurdles to realizing the full value of these resources. For example, the identification of novel molecular targets for therapies through GWAS information presupposes the understanding of the specific mechanism associated with an identified genetic association. However, genetic associations described through GWAS studies are not able to provide more than an association estimate as a proxy to causality. Secondly, the lack of integration with additional genetic information maps (such as expression or protein QTL data) makes them unable to predict the mechanism of action (MoA) of a particular gene to a phenotype, information that is fundamental for the development of a therapeutic program. Thirdly, most GWAS have a narrow focus on traits, which is often incomplete compared to the overall set of traits relevant for understanding of a potential drug-target landscape. Unfortunately, the capacity to conduct large-scale phenotypic exploration of genotypic associations, which could overcome some of these limitations, are available at scale to very few biobanks, and still face considerable challenges in terms of power and harmonization with other domains of biological understanding.

To overcome some of these limitations, one would ideally need to use orthogonal sources of evidence, such as causal evidence from Mendelian Randomization (MR), association with rare protein-coding variants, known causal associations between genes and Mendelian disorders, insights form the human protein-protein interactome, and animal genetics, all of which are exposed to different biases. In fact, in recent years several groups have attempted such strategies, usually focusing on a single, or a few, trait(s)^10^.

However, these initiatives have also faced significant challenges. First, they require trait/phenotype harmonization between sources of biological information as different as drug databases, clinical entities, rare genetic syndromes, and even animal phenotypes. Second, although previous efforts have quantified the “odds of success” of future programs based on historical data, this approach tends to be conducted highlighting the enrichment between genetic association between genes and phenotypes and its over-representation in different dimensions of biological knowledge but without deriving a classification metric that could allow for the use of multiple sources of evidence within the same model.

We used causal inference methods^11,12^, associated with a large list of genetic instruments, to address some of these shortcomings^13^. In addition, the use of catalogs describing the associations between genetic variants and thousands of human traits provides a resource for the identification and validation of novel targets^14^. Finally, we propose that the integration of these results for a large number of traits provides an emerging property of this approach by highlighting common mechanisms between medical entities previously considered distinct.

To overcome these limitations, we meta-analyzed GWAS data from the Million Veteran Program^15^ (MVP), the UK Biobank, and FinnGen covering more than 1 million individuals and 1,885 traits harmonized through state of the art algorithms, considerable improvement in sample size and number of phenotypes compared to similar attempts. Then, we leverage this resource with three GWAS-pQTLs using Somascan V4 (5K proteins) and two GWAS-eQTL resources to conduct a genome-wide MR-PheWAS. These resources were then combined with orthogonal sources enabled by novel methods on phenotype harmonization, to generate a machine learning (ML) model to quantify the odds of success of a potential target on a given disease using historical data on approved drugs. Finally, we applied our ML model to 69K gene-trait associations to prioritize novel drug targets, identify indication expansion for investigational drugs, and drug repurposing opportunities for licensed drugs with higher odds of success. We believe the resources made available by this work can have broad use in drug discovery programs.

## Results

### General findings from a large-scale two-sample MR exploration on human disease

In this effort, we systematically studied 2,011 (2,003 after excluding height related traits) distinct diseases/biological traits meta-analyzed from MVP, UK Biobank, and FinnGen encompassing summary statistics derived from more than 1,200,000 individuals (**Supplementary Table 1**). We analyzed each phenotype against 15,919 genes, instrumented through 15,700 transcripts (eQTLs derived from GTEx and eQTLGen) and 1,933 protein (pQTLs derived from ARIC, deCODE, and Fenland) levels through two-sample Mendelian Randomization (MR). In total, we tested 31,525,236 unique gene/protein-trait associations (**Figure 1**). A detailed descriptive analysis of the genetic instruments used is available as **Supplementary Appendix 1**.

**FIGURE 1.**
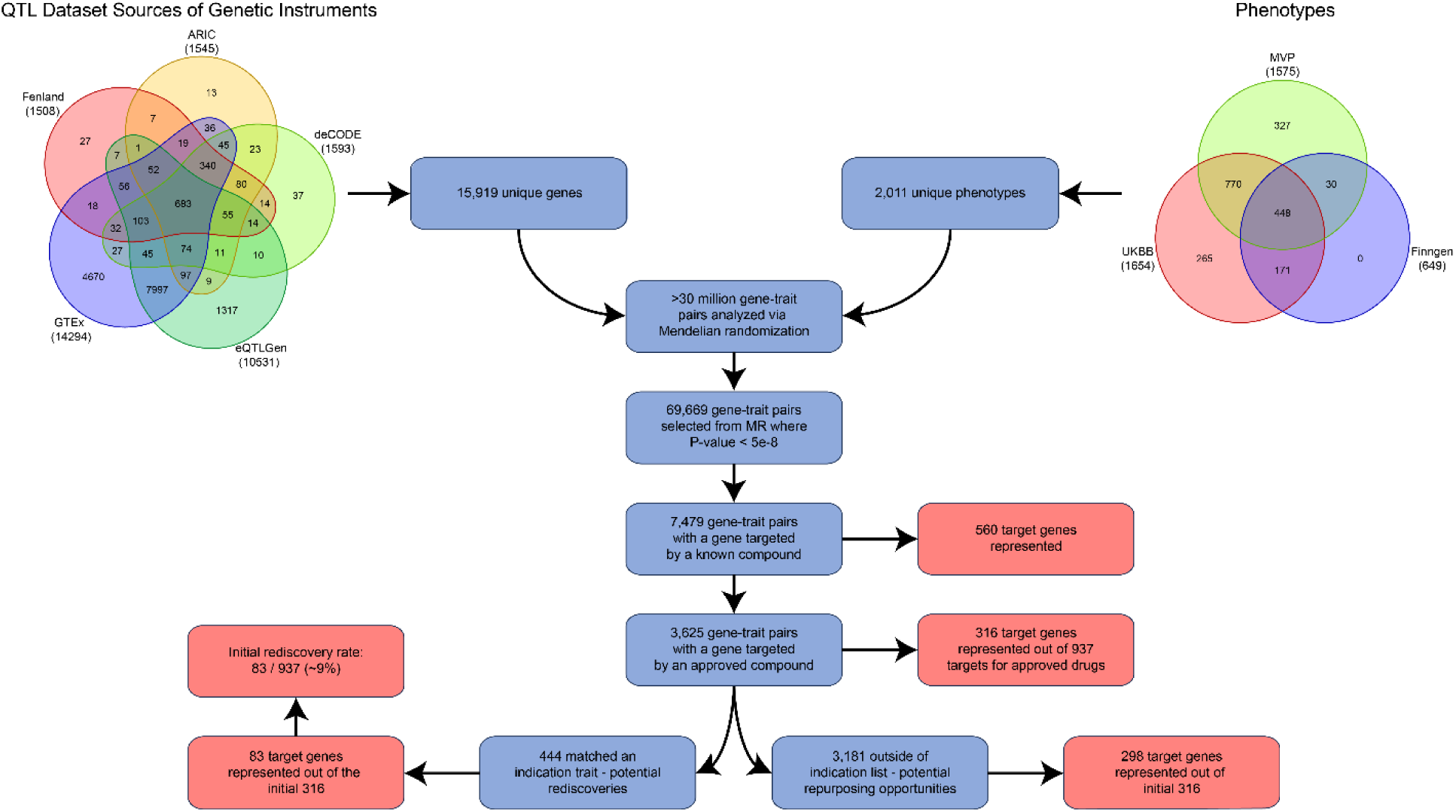
– A flowchart demonstrating the main findings from the pipeline and the resulting counts of both rediscovery and repurposing opportunities.

From the two-sample MR analysis results, we observed 108,107 gene-trait pairs with a p-value ≤ 1.6e-9 (79,114 unique gene-traits). If the same transcript/protein was identified as potentially causal of the same trait using more than one instrument source, we compared whether the predicted directionality of effect was concordant among the sources (i.e., among different eQTL, including different GTEx tissues; or among different pQTL sources). If there was discordance, these gene-trait pairs were removed from consideration resulting in 69,669 unique gene-trait pairs (Supplementary File 1, Supplementary Figure 1) (from now on referred as selected MR results) selected for downstream analysis (more details in online methods).

We systematically compared our results to those available in the GWAS Catalog (as of Jan/2023). Of the 69,669 selected results, 42,737 (∼61%) corresponded, in the GWAS Catalog, to a previously reported genome-wide significant association within the region (see methods) assigned to the same trait. We observed 26,932 (∼39%) causal effects that were considered novel. That is, there was no significant result described in the GWAS Catalog for the same trait that mapped to the same genomic region. When considering traits mapped to a similar parental disease hierarchy, as opposed to the same trait, 8,932 (∼13%) of our selected results were considered novel for they did not map to any association described in the GWAS Catalog for any trait in the same disease hierarchy. The described degree of novelty when compared to available gene-trait association in the GWAS Catalog, together with the fact that MR explicitly instruments the relationship between a gene and a trait, highlight the advantages of our approach over a GWAS results-based approach.

### Triangulation of MR results with orthogonal sources to annotate novel findings

To further increase the confidence on our MR findings, we triangulate our MR hits with orthogonal sources such as OMIM^16^, ClinVar^15^, pLOF burden analysis of rare variants from the UK Biobank^17^, and the MGI mouse knock-down database^18^ for gene-trait pairs mapped to the gene-trait pairs tested in our study. To enable triangulation at scale of MR hits with orthogonal sources, systematic mapping of traits across all these sources is essential. Whereas mapping the genes is straightforward, mapping phenotypes among these databases and our study is more complex. To map phenotypes among these databases, we created for each trait, in each of the datasets, three different mapping schemes with the traits from the list of phenotypes we used in the MR analysis. These were: an exact match (same trait name or concept), and a parent term match (where we mapped all the used traits to one or multiple parent terms pre-defined in a system level dictionary), as used for mapping novelty in the set of selected MR results (see section above). In addition, we derived a semantic distance match (where terms were matched by a semantic distance approach using embeddings of all traits present in all used databases). We discuss in detail our phenotype mapping strategy in the Supplementary Methods section.

We mapped all tested gene-trait pairs in MR to the curated catalogs of orthogonal biological information. Selected MR gene-traits were significantly more likely to be represented by all sources of biological information as compared to non-selected MR gene-trait pairs (**Figure 2**). This was observed for all schemes used for mapping phenotypes between datasets. For example, if the gene being tested for a specific trait has been previously described as a gene-trait association in OMIM, there is a 68-fold increased odds of the gene-trait being a selected MR result (p-value = 2.77e-05). Similar behaviors are observed if the gene-trait being tested has been identified in a pLOF rare variant burden analysis on the UK Biobank (OR = 86, p-value = 9.76e-62), or described as a gene causing the trait in ClinVar (OR = 4.00, p-value = 1.97e-04). Interestingly, enrichment for KO models was lower than anticipated (OR=2.02 and OR=1.46 depending on the phenotype mapping strategy) (Supplementary Table 2 and **Figure 2**). Of note, the four described orthogonal sources were significantly independent. That is, they were all significantly associated with a selected MR gene-trait in a multivariable model adjusting for all orthogonal sources (Supplementary Table 3).

**FIGURE 2.**
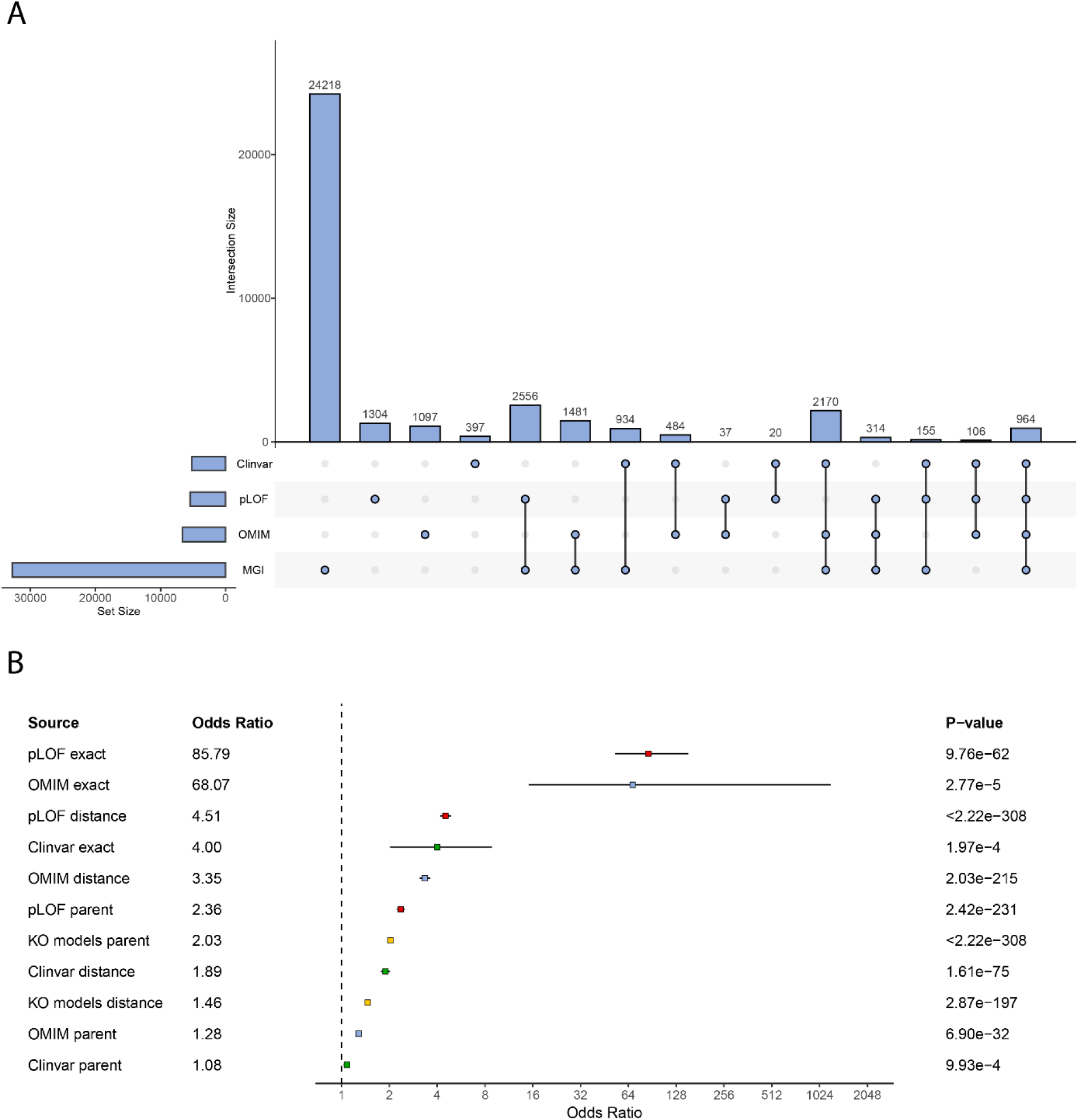
– **A.** Upset plot representing the different intersection numbers of gene-traits among the used sources of orthogonal biological information. **B**. Forest plot of the association between the different biological features as predictors of being a significant MR gene-trait. Different distance metrics used for capturing a trait match were exact = same EFO term for both MVP and biological database; distant = closest 3% terms in the ranked list between MVP and biological database using a semantic distance metric; parent = same parent term in both MVP and biological database.

Although these findings reinforce a putative causal role of the selected gene-traits, from a drug discovery translational perspective it would be ideal to quantify the odds of a selected gene-trait to be developed into a successful drug target. To answer this question, we used historical data from successful drugs with known efficacy targets.

### Mapping causal genes and traits to approved drugs and their indications

Using ChEMBL^19^ release 34 we mapped all selected gene-traits from our MR effort to existing drug targets and their indications. To this end, we have systematically mapped all drug targets described in ChEMBL 34 (1,533 mapped targets from 4,315 in clinical-development or approved drugs) and disease indications (2,031 unique indications) into 33,102 unique drug target-indication pairs (Supplementary File 2). From the 6,447 set of unique genes in the selected MR gene-trait results (69,669 gene-trait pairs described above), 560 were the target of an existing compound, including preclinical compounds (represented in 7,479 selected gene-traits) (Supplementary Table 4). From these, 58 genes mapped to the target of an approved drug. This set corresponds to 6% of all described targets for approved drugs in ChEMBL 34 (a total of 937 approved drug targets described). Genes that are targets of drugs in development (listed as developmental phase I to III) are significantly over-represented in our set of selected MR gene-traits as compared to genes that are not targets of an existing drug (Supplementary Table 4). Looking at potential targets that are currently listed specifically in Phase 3 clinical development by ChEMBL 34, from the 7,479 selected gene-traits that target a drug in clinical development, 1,477 had their maximal phase listed as 3. From these, we were able to identify 123 selected gene-traits where the indication of the ongoing trial efforts matches the trait the gene was associated to in our MR results (Supplementary Table 5, Supplementary Figure 2).

From the 3,625 gene-traits with genes that were targets of approved drugs, we mapped 257 selected gene-traits (70 unique indications) that fully recapitulated both the approved drug target and its approved indication. We consider these gene-trait rediscoveries, and they represent 58 unique drug targets (6% of all approved drugs targets) (**Figure 3**). Using the MR estimate and the approved drug mechanism of action (MoA), we compared within the set of rediscoveries whether the calculated MR estimate direction of effect was in agreement with what would be predicted by considering the approved drug MoA. The MR estimated direction of effect correctly predicted the expected MoA 84% of the time (p-value = 4.46e-09, 95% CI 74% – 92%). We did not observe an association between being identified through a pQTL and a higher odds of correctly predicting the expected MoA (p=0.47, odds of correctly predicting MoA for eQTLs 84%, odds of correctly predicting MoA for pQTLs 89%).

**FIGURE 3.**
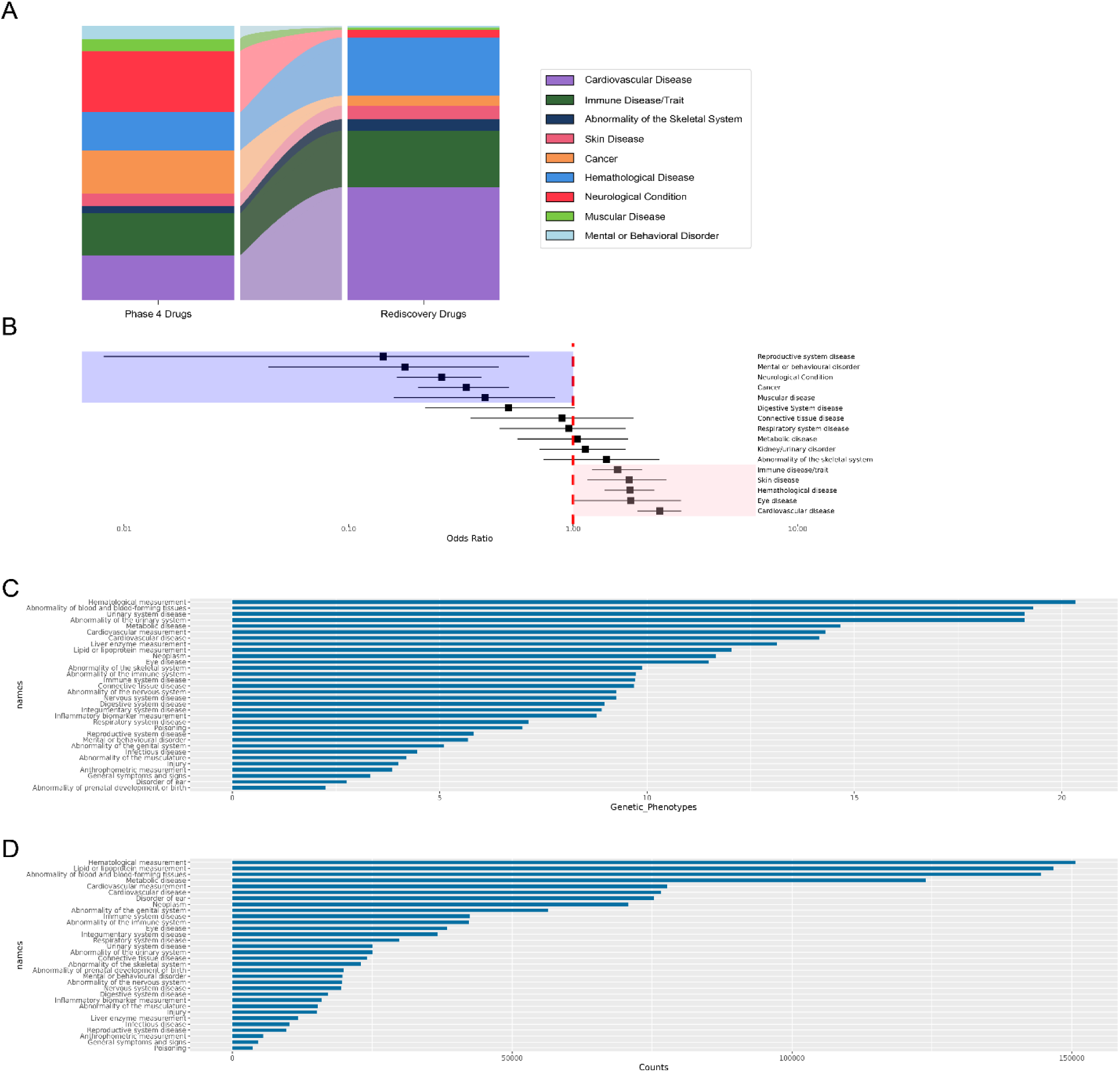
– Results from genetic rediscoveries of approved drugs. Panel A, stacked bar plot of the distribution of parent terms for all currently approved drugs indications (left bar), and distribution of parent terms among gene-trait pairs considered rediscoveries (right bar). Panel B, forest plot representing the over or under-representation of specific parent terms among rediscoveries. Panel C, mean number of available genetic phenotypes per parent term indication. Panel D, mean number of cases used to calculate the association between genetic variants and the outcome per parent term.

Rediscoveries were notable for a range of different diseases and organ systems (Supplementary Table 6). Nonetheless, by comparing the drug indications using a systems-based approach we observed that drugs targeting some biological systems were over-represented as compared to the distribution of all approved drugs and their indications. Specifically, drugs targeting cardiovascular system diseases (OR = 3.3, p-value = 2.88e-13) were more likely to be rediscovered through our approach, whereas drugs targeting different types of cancer (OR = 0.2, p-value = 5.31e-04) or neurological conditions (OR = 0.1, p-value = 1.22e-05), among others, were less likely to be rediscovered (**Figure 3**, panels B and C). We explored technical variables that could explain a higher or lower rediscovery rate of drugs targeting specific organ systems such as the number of mapped phenotypes between GWAS data and approved drug indications, the total number of approved indications for a particular organ system, and predictors associated with the statistical power of available GWAS (such as the total number of cases). The most important predictor was the percentage of mapped phenotypes with less than 10,000 cases (p-value = 0.003), suggesting that the under-representation of rediscoveries for some disease types might be mitigated by increasing the number of GWAS studies (and, consequently, the number of cases) used in the MR analysis for some specific diseases such as cancers, neurological and psychiatric conditions in future iterations of this work (Supplementary Table 7).

### Novel potential repurposing opportunities observed for approved drug targets

A corollary of the mapping of selected MR gene-traits to approved drug targets is the possibility to unravel drug repurposing opportunities. These are the selected gene-trait pairs mapped to a gene that is the target of an approved drug, but not associated to a trait in the listed approved indications for that drug. Rather, the associated trait is a novel one putatively modulated by the approved drug. Of the 3,625 selected gene-traits that mapped to an approved drug target, 3,366 had a trait different from the one described as an approved indication in ChEMBL34. These encompass 1,589 different drugs and 960 unique novel potential repurposing (354 indications) or safety (236 indications) concerns for currently approved drugs. In **Figure 4** we show the transition between approved indication disease systems and repurposing disease systems. A complete list of all observed potential repurposing opportunities can be found in Supplementary Table 8.

**FIGURE 4.**
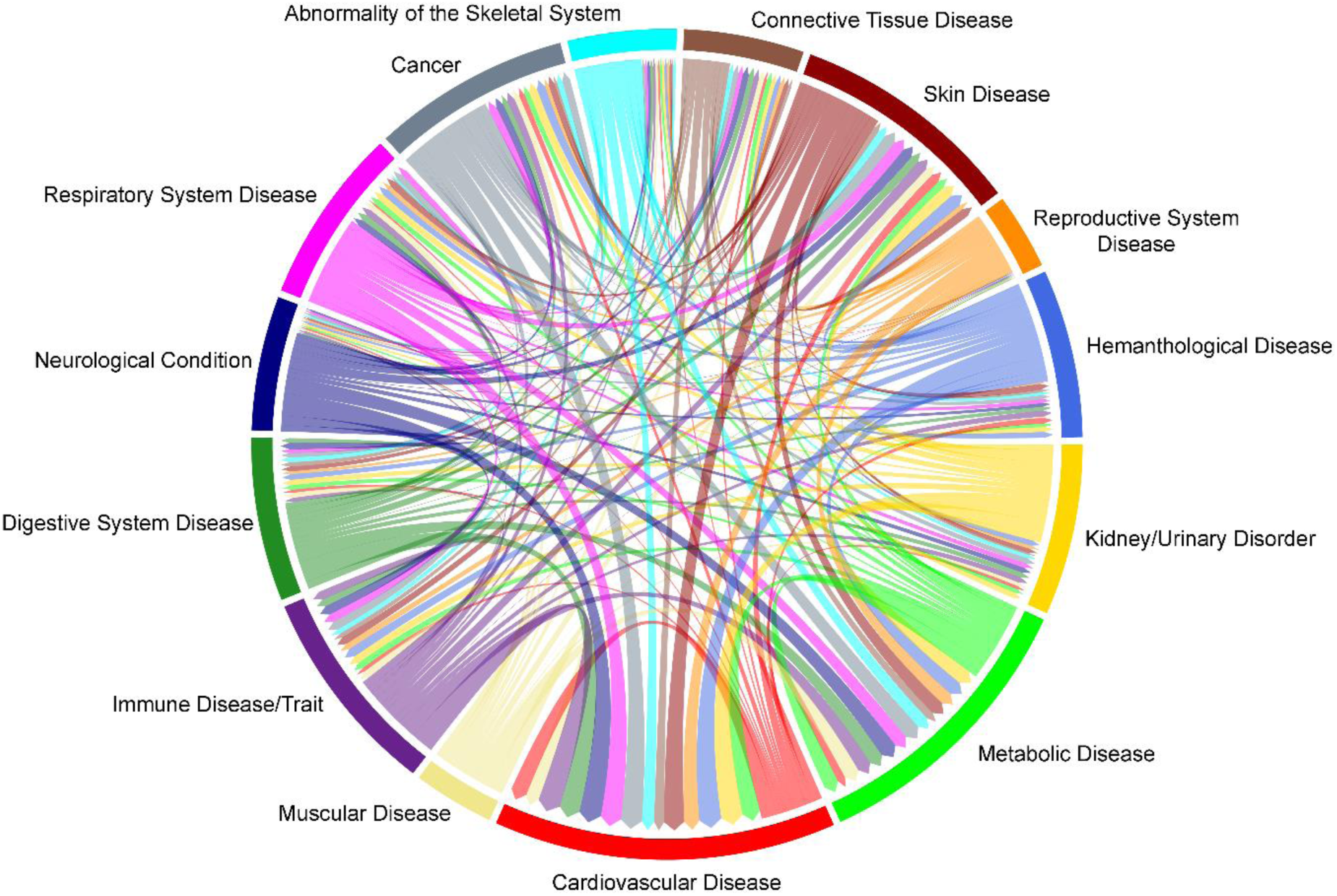
– Circle plot representing the flow of disease categories for approved drug indications to the disease categories of potential repurposing opportunities based upon our findings.

Among the several observed repurposing opportunities, some might deserve special consideration due to the prevalence of the novel indication and/or the large usage of the approved drug: Probucol, an ABCA1 inhibitor with lipid-lowering properties for glaucoma; Galcanezumab, a CALCB inhibitor approved for migraine profilaxis for obesity; Anrukinzumab, an IL13 antagonist approved for ulcerative colitis for psoriasis; Tocilizumab, an IL6R antagonist approved for rheumatoid arthritis for atrial fibrillation;; Metformin, a NADH dehydrogenase inhibitor approved for type 2 diabetes for atrial fibrillation; among many others.

It is important to note, however, that these initial repurposing opportunities should be contemplated considering the potential mis-assignment of directionality given by blood-based eQTLs and pQTLs^20^ and a comprehensive examination of safety concerns is beyond the scope of the present work. One example that can be highlighted from our results is the presence of Trastuzumab, an ERBB2 inhibitor, as a potential repurposing opportunity for both atrial fibrillation and heart failure (Supplementary Table 8). Although there is the possibility that Trastuzumab might be protective against both conditions in specific scenarios, clinical knowledge highly suggests the contrary. Trastuzumab is a known cardiotoxic compound^21^ and most probably the highlighted signal should be classified as a safety concern rather than a repurposing opportunity.

### Selected MR gene-trait pairs are predictors of approved drug indications

A mapping framework that triangulates data from four key areas—gene targets of existing pharmaceuticals, their approved therapeutic uses, results from genome-wide association studies (GWAS), and available genetic proxies of molecular exposures—can be employed. This framework serves as a foundation for developing and validating predictive models for Mendelian Randomization (MR) analyses of gene-trait associations using the gene targets and indications of currently approved drugs. With this in mind, we have mapped approved drug targets to unique clinical indications (to reduce bias due to similar clinical indications for the same drug target); mapped approved indications to the phenotypes with available genetic association summary statistics; and created a benchmark file containing positive and negative controls for training a model for approved drug indications. Negative controls were paired to positive controls by the clinical indication (to avoid bias due to increased GWAS statistical power for commonly approved indications) and derived by randomly selecting a gene from the gene set with at least one significant genome-wide MR results (to avoid the bias of creating negative gene-trait pairs using genes with weak genetic instruments). For each existing positive-control we created 10 random negative-controls. Our training dataset was composed of 4,027 positive controls and 40,216 negative controls. Our results show that an MR result with robust evidence of a causal effect is associated with a 2.77 (95% CI: 2.3 – 3.4) higher odds of the associated gene-trait being a positive control (that is, one of an approved drug-indication) (p-value = <2e-16). This estimate is concordant to previously published ones using a smaller number of phenotypes^10,22–25^. Acknowledging that other criteria can be used to select gene-trait pairs for further analysis, and that depending on the used criteria the sensitivity and specificity of the approach can significantly change, we provide in the Supplementary Appendix 2 a comparison of different selection criteria and the resultant number of selected gene-trait pairs, rediscoveries, and relative odds of a selected gene-trait to be an approved drug target and indication.

### Genetic colocalization between GWAS and instrument signals improves specificity and the positive predictive value at the cost of reducing sensitivity for detecting approved drug targets and their indications

Prior work on the use of MR for early target identification has used colocalization as a filtering step after the initial MR result^26,27^. To assess the use of genetic colocalization within this strategy, we evaluated the impact of using colocalization for filtering selected MR results (that is, accepting as potential new targets those that have an MR p-value < 1.6e-9 and also significantly show a high colocalization signal, defined as selected MR results with at least one PPH4 region (both traits share a single causal variant within a genomic window defined around an instrumented variant) colocalization signal > 0.8 per instrumented gene/protein.

As expected, using genetic colocalization as a filter reduced the number of selected MR results from the initial 69,669 gene-trait pairs to 54,636 (78%) gene-trait pairs when at least one significant PPH4 result was required.

The consequence of applying this filter was a reduced number of rediscoveries, but an increase in the enrichment associated with a selected gene-trait. While gene-traits only passing MR were associated with an OR = 2.77 (95% CI: 2.3 – 3.4)) of being an approved drug indication, restricting selected MR results to only those passing at least one colocalization test defined a set of gene-traits associated with a 3.09-fold enrichment (95% CI: 2.5 – 3.8, p-value = <2e-16). The price paid by this increased positive predictive value was a reduced sensitivity: from an initial set of 148 selected gene-traits rediscoveries unique indications, 109 were observed when requiring at least one colocalization result.

With these results we posit that the reduced sensitivity could significantly impact the potential for new discoveries. Therefore, we decided to not use genetic colocalization as a required criteria for downstream analysis of selected MR results, but rather as a potential predictor of a selected gene-trait to be a true positive (see section on training a classifier for true-positive associations).

### Orthogonal information is predictive of approved drug targets and indications

Using the same strategy to show the capacity of a selected MR result to predict an approved drug target-indication, we inquired whether the trait mapping schemes and secondary sources of biological information we previously described as associated with a selected MR gene-traits, could also inform on the odds of a particular gene-trait being the target of an approved drug indication. Gene-traits that could be mapped to any of the different biological databases (and in any of the three trait mapping schemes used) were significantly more likely to be approved drug targets and indication (Supplementary Table 9).

### Understanding MR-identified target landscape with protein interaction networks

There has been interest in the potential to extend the space of credible drug targets by examining proteins that interact with “genetically validated” targets. Such targets may be preferred to the original because they have improved properties, e.g., with respect to safety or chemical tractability^28^. Here we tested whether information from the protein interactome of a selected MR gene-trait pair could inform the predictive value of it being associated with an approved drug for the same gene-trait pair. This was done by an expansion and modification of a similar approach used to define the enrichment of MR results within the set of approved drug-indications^29^. Specifically, we constructed aggregated protein-protein interactions network containing seven different protein-protein interaction (PPI) resources: Complex^29^, Lit BM^30,31^, Metabase (https://portal.genego.com/), OmniPath^32^, HIPPIE^33–36^, HI union^30^, and STRING^37^ (details on pre-processing and assembly can be found on Supplementary methods). Using the aggregated network, we observed that the number of first-degree protein partners was higher in selected MR gene-traits as compared to gene-traits that did not reach statistical significance (p-value < 2e-16). More importantly, the PPI information was predictive of associations between a gene-trait pair and an approved drug for the trait. Namely, having a significant MR result and being a gene that is a PPI with another gene that also has a significant MR result for the same trait increases the odds of the queried gene-trait be an approved drug target and indication [OR = 1.89 (95% CI: 1.8 – 2.0, p-value = < 2e-16)]. In addition, the odds of a given gene-trait pair being associated with an approved drug was significantly higher [OR = 1.17 (95% CI: 1.1 – 1.3, p-value = 1.5e-05)], when a first-degree PPI of the gene was associated with an approved drug for the same trait compared to when no PPIs of the gene were approved drug targets for the trait. Interestingly, the different PPI resources did not share a large amount of specific protein-protein interactions (see Supplementary Figure 3) and, hence, their value to the overall enrichment was mostly additive (please see Supplementary Table 10 for a fully adjusted model). Finally, although having a first-degree PPI of the gene associated with an approved drug for the same trait, expectedly, positively associated with higher odds of the gene-trait being mapped to an approved drug indication in most used PPI resources, in HI union, a PPI resource derived mostly from cancer cell lines, unexpectedly, this occurrence was significantly and independently associated with a reduced odds of the initial gene-trait being mapped to an approved drug indication.

A second strategy that we devised aiming at using information provided by PPI was to determine whether a first-degree partner of a selected MR gene-trait was also a selected MR gene-trait to the same trait and whether this information influenced the odds of the initial gene-trait to be mapped to an approved drug-indication. For this analysis we recursively tested more than 19 million gene-trait combinations and calculated the number of times a first-degree protein partner had robust causal evidence to the same trait of a given gene-trait pair. Results showed that selected MR gene-trait pairs were enriched for the presence of a first-degree protein partner also with significant MR results for the same trait, independently of the total number of interacting proteins of the gene. Having a first-degree protein partner with a significant MR result for the same trait significantly increased the odds of being a selected MR result by 22-fold (p-value < 2e-16). In addition, similarly to the discussion above in the case of protein partners being a target of an approved drug, for a given gene-trait pair, having protein partners associated with the same trait also significantly increased the odds of the initial pair being the target and indication of an approved drug. The larger the number of protein partners associated with the trait of a given gene-trait pair, the higher the odds of the gene-trait pair being the target of an approved drug indication (for example, each added protein partner associated with the same trait increases in 4% the odds of the initial gene-trait to be an approved drug indication, p-value <2e-16). This reinforces the intuition that convergence of multiple genetic associations on a pathway identifies key disease-causing pathways.

### Ranking selected MR results by the probability of success in drug development

From the 69,669 unique gene-trait pairs identified by MR only a very small number were considered rediscoveries (i.e., a selected gene-trait pair that can confidently be labelled as a true positive finding). The great majority of selected gene-trait pairs here described are, although potentially interesting, still suggestive, at the best, of a viable drug target and clinical indication. Leveraging the previously described resources and phenotype mapping strategies, we used the approved drug indications in ChEMBL 34 to derive a classifier able to predict the likelihood of a given gene-trait to be developed into an approved drug indication. For this, we used a machine learning model approach and engineered 40 features based on the different enrichments we described between a selected MR result and approved drug targets and indications (details of each engineered feature can be found at Supplementary Table 11). We describe the modelling approach in detail in the Supplementary Methods section. Briefly, our classifier was trained using 80% of the entries in the previously described benchmark dataset and tested in the remaining 20% which was kept as a hold-out set only used to test the final model. We selected the most influential features and integrated these into a classifier for approved drug indications. The overall accuracy (**Figure 5**) was observed in a precision-recall AUC of 0.79. Additional characteristics of the model performances can be found at the Supplementary Appendix 2 and Supplementary Table 11.

**FIGURE 5.**
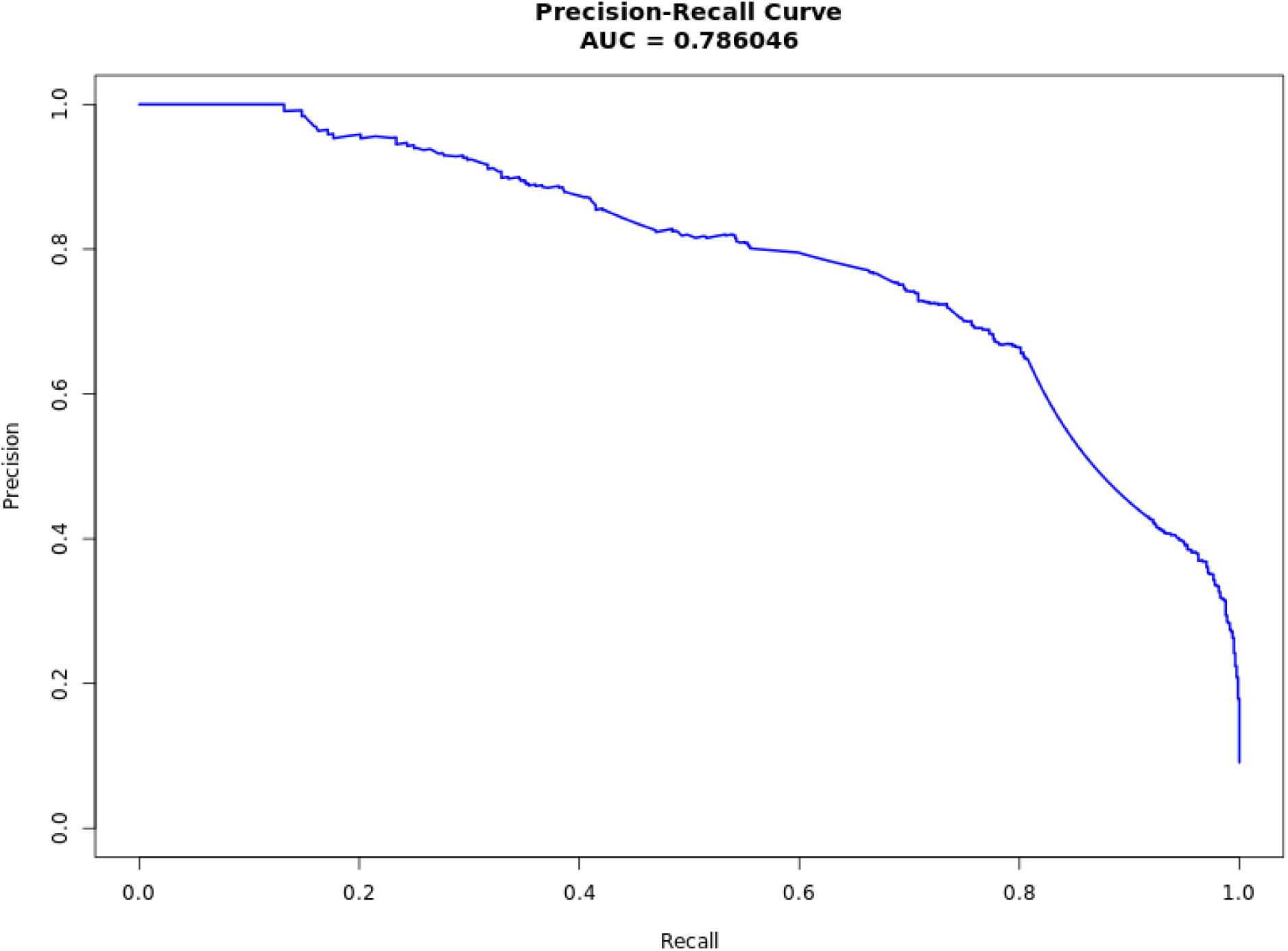
– Precision recall estimate of our classifier.

Using this model, we derived predicted probabilities for all 69,669 selected gene-trait pairs (Supplementary File 1). Finally, we ranked all selected gene-trait pairs based on the probability described by this model. This type of data representation is now able to be used to compare selected MR gene-trait pairs regarding their similarity to gene-trait pairs of approved drug targets and indications.

The predicted probabilities derived from our model provide the possibility to be more restrictive when selecting MR results for downstream exploration. For example, using a very restrictive cut-off probability in which both the positive and negative predictive values are above 0.8, we still have 338 gene-traits. (Supplementary Table 4).

### Prioritization of targets for drug development identify ANXA2 as a novel target for dyslipidemia

Among the many uses of the developed framework and models, one is their utilization in prioritizing new drug development programs^38^. In fact, the probabilistic nature and output of our classifier permits that, among different pre-existing gene-trait candidates, one can identify the most promising lead to an approved drug indication. Here we exemplify this approach describing our results for anti-lipidemic and anti-obesity targets.

Using ChEMBL 34 we identified 9 approved targets for treating dyslipidemia in humans. From these, our MR approach was able to rediscover three targets (*HMGCR*, *PCSK9*, *NPC1L1*). We also identified 11 targets among the 36 targets that were, or are being, tested for dyslipidemia (that is, are below phase IV, but in clinical testing phase 1-3). Additionally, using 25 different lipid traits that are part of our effort, we observed 4,472 selected MR results (892 unique genes associated with at least one lipid-related trait). Among 1,236 genes previously annotated to be associated with a lipids GWAS hit^39^, we identified 291 (33%) in our list of selected MR results also associated with lipids. From the protein-coding genes with rare variants previously associated with lipid traits (121), we were able to recover 81 (67%) of the total described. Considering all previously associated genes from both GWAS and rare variant analysis, and assuming a genomic window of 100Kb around each gene, we defined 750 unique genomic loci with at least one previously described lipid gene. Our results were able to map at least one MR associated gene to 291 of these loci (39%). From the 291 we were able to map at least one of the previously mapped genes in 199 instances. Interestingly, in 136 of previously mapped lipids loci, our results suggest at least one other gene as the causal effector of inter-individual differences in lipid levels in these genomic intervals (Supplementary Table 12). In addition, for loci where several genes were mapped as potentially causal for regulating lipid levels, the use of our ranking model was able to suggest the best candidate amongst associated genes. For example, on chr14:91865991-92140896, previous GWAS observation pointed towards *TRIP11* as a lipids-related gene ^40,41^. We did not find causal evidence for *TRIP11*. Nonetheless, *ATXN3* was an MR hit with robust causal evidence in the same genomic interval. Our ranking model classified *ATXN3* as a gene with a high probability of being a drug target for lipids. From the selected MR results associated with lipids that were neither previously described, nor mapped to one of the previously described loci, we have identified 369 novel genes. Tuning our ranking model to a positive predictive value of 0.70, we identified 7 novel genes with high probability of being targets for drugs modulating lipid levels: *ANXA2, TNF, HAVCR2, MPIG6B, HIST1H1C, TMEM57, GSDMC*. Of particular interest is the presence of Annexin A2 (ANXA2) in the list, a natural inhibitor of PCSK9 and an endogenous regulator of LDLR degradation^42,43^.

Taken together the evidence supports the results of our MR analysis and predictive model and suggests that our data may disclose several novel therapeutic targets for a wide range of human disorders.

## Discussion

Here, we have systematically leveraged genetic association results from a large number of phenotypes and integrated different database sources to generate almost 70,000 potential causal effects of genes/proteins on disease-relevant traits. This endeavor extends previous efforts of using genetic information for early therapeutic target identification by almost 10-fold^44–47^ and provides a new resource for investigation of the polygenic causes of a wide range of human phenotypes. By increasing the breadth of our gene/phenotype matrix of causal effects, we allow for a more diverse exploration of human disease diversity, as well as the therapeutic opportunities that might be revealed by using empirical population genetic evidence.

Initially, it is imperative to highlight the importance and challenges of highly diverse large-scale biobanks and data sources integration^48^. This was one of the main challenges of the current work. In fact, there are significant challenges associated with the mapping and integration of large-scale biobanks developed in different medical systems (MVP, UK Biobank and FinnGen), genetic instrument resources derived from different research programs (GTEx, eQTLGen, ARIC, Fenland, and deCODE studies); different bioinformatic databases (OMIM, MGI, ClinVar); different sources of experimental molecular findings (PPI networks from different studies). For this, we have approached each biological data source as a different construct and proposed different mapping schemes at the molecular and phenotype levels. While at the molecular level we have opted for a gene-based approach, which is facilitated by using a Mendelian Randomization framework; at the phenotype level, mappings can be more nuanced and difficult. We here opted for a hybrid phenotype mapping approach that used both the specificity of ontology-based classifications (such as EFO terms), but also the more fluid capacity that medical term embeddings allow for. On top of this, we have been able to manually curate all different disease/trait-associated databases and map each of their terms to pre-defined parental terms (see Supplementary Table 13) that allowed for more general disease classifications. The challenges posed by phenotype harmonization and annotations were paramount, given the disparate data products and annotation schemes in biology, not to mention the variations in model organisms and their associated phenotypes. Despite the availability of ontologies such as SNOMED, FHIR, and the ICD coding system, which are increasingly utilized in medicine, they fall short in addressing the complex needs of our study—mapping different sources to a common denominator and understanding the phenotypic distances within and across sources. Our use of a text embedding vector store, constructed through the text vector representation of disease names using a semantic similarity algorithm, emerged as a pragmatic solution. This approach successfully bridged the gap between the disease name/concept and higher structured disease hierarchies such as organ systems. Furthermore, the association estimates between the different distance metrics aligned with our expectations, validating the efficacy of our methodology.

Although several prior tentative methods of integrating genetic, animal models, and molecular interaction data, coupled with the sophisticated mapping of phenotype hierarchies, have been used in specific biological and medical contexts^49–53^, we here provide an approach that is anchored by large-scale population genetic evidence. The links created in our work are mostly based on a gene/protein level causal effects defined by Mendelian Randomization. This approach relies on the duality defined by, at one side, the reliance on accurate genetic association statistics for the studied phenotype; on the other, the capacity to capture unbiased interindividual variability on genetically determined exposures. Our approach has original contributions on both sides. On the outcome side, we are querying genetic association data from more than 2,000 different human phenotypes. Sample size and statistical power were maximized by, whenever possible, meta-analyzing the largest genetic biobank studies currently available: the UK Biobank, the FinnGen Study, and the Million Veteran Program. This effort is reflected in the large diversity of available phenotypes for analysis and on the mean number of cases per phenotype analyzed. On the exposure side, we have used 5 different sources of genetic instruments, capturing both transcript and protein levels for many molecular entities. In addition, our instrument selection process only selected conditionally independent *cis* instruments, thus reducing the chances of bias due to both model misspecification and horizontal pleiotropy. Nonetheless, it is worth noting that horizontal pleiotropy is difficult to avoid for different genes/proteins (particularly when overlapping or in proximity) which can be regulated by a common subset of genetic instruments or confounded by linkage disequilibrium. This makes the dissection at this level particularly challenging, especially for an effort conducted at the present scale. Despite this limitation, the tunneling of our inferences at the gene level, as opposed to at the variant level, which is usually the case for GWAS or PheWAS efforts, has the advantage of reducing the dimensionality of the results dataset and to add an extra layer of support for target identification.

Instead of focusing our inference to only transcripts or proteins, we decided to use several independent genetic instrument sources and contemplated the challenge of making inferences for the entire set of protein coding genes in the human genome. Notably, there was very high correlation between the estimate of effects within the three pQTL sources, and within the two eQTL sources, despite the use of different genetic instruments among the different sources (Supplementary Figure 5). The correlation of predicted effects between eQTLs and pQTLs were, expectedly, smaller. Despite being comprehensive, this approach has clear limitations due to all the situations in which transcripts and proteins operate in distinct directionalities. As a result, it is still hard to define a particular effect that should be emulated for in a new drug. A cautionary tale is that, although the predicted directionality of effect was correctly predicted in approximately 85% of the instances, in about 15% we have significantly identified the correct gene-trait pair, but our results pointed towards the contrary direction. A direct comparison between the different sources of genetic instruments was beyond the objectives of the present work and might prove challenging because of the very different nature of the study designs, technologies, statistical power, and biological relevance of each source. We believe this is a fruitful new avenue of research that should be explored in future analyses and provide the results that would be observed by our approach if we used only eQTL or pQTL sources (Supplementary Appendix 3). It is also important to note that powerful new sets of genetic instruments for molecular phenotypes are being developed such as the novel UK Biobank proteomics dataset.

Drug rediscovery, in this manuscript, means identifying a gene-trait association that corresponds to one of the known efficacy targets of an approved drug (as the gene) and is linked to the same medical indication or therapeutic use for which the drug was originally approved (as the trait). Essentially, it is discovering genetic evidence that supports the target and indication of a drug^54^. We were able to rediscover approximately 6% of known approved drug targets in ChEMBL 34 (Supplementary Table 6). Some drug indications were rediscovered several times, that is in several different tested phenotypes, and about half of all rediscoveries have not been previously described as a gene-trait associated in a previous GWAS study, that is, where the drug indication was previously supported by genetic evidence from common variants genetic association. As such, the concept of rediscovery reaches beyond the replication of previous genetic associations of genes and phenotypes that are drug targets and trait indications. These facts highlight both the power of the approach and the potential to still be leveraged in its future iterations. Of particular importance, because we have benchmarked the different steps of our procedure using a list of curated approved drug indications, we can use this information to guide new methods, data extraction and needs of the current pipeline so that new iterations can be more sensitive and specific. This might be exemplified in the set of technical predictors of rediscoveries. We showed that for some specific disease categories, the number of explored phenotypes and case counts are still bottlenecks that need to be surpassed. Another feature of note is the capacity to use continuous variables as proxies for disease/drug indications. For example, most of the enrichment in the rediscovery rate of cardiovascular drugs can be traced to anti-lipidemic phenotypes that can be captured by lipid measurements, continuous variables available in all three used biobanks. Finally, further investigation is needed to determine whether the same genetic information used to establish significant Mendelian Randomization effects can also be employed to stratify individuals based on the varying effectiveness of drugs identified through rediscovery signals.

Building on the insights gleaned from the analysis of 69,669 unique gene-trait pairs identified, our approach has effectively distinguished a small subset of rediscoveries, affirming their status as true positive findings. Nonetheless, most significant gene-trait pairs, though intriguing and potentially indicative of viable drug targets and clinical indications, are still in a preliminary stage. Leveraging the comprehensive resources and phenotype mapping strategies previously outlined, we have crafted a classifier, utilizing a list of approved drug indications from ChEMBL 34, to predict the likelihood of a given gene-trait pair progressing to an approved drug indication. Through the application of XGBoost the classifier exhibited a precision-recall AUC of 0.79. At this point, it is important to highlight the conceptual limitation of our model, which is maximizing the selection of gene-traits that have a similar feature profile to that of approved drugs and their indications. Factors modulating a successful drug program are determined by many more factors than only the causal association between a gene product and a disease, and this should be taken into consideration when interpreting our results. In addition, some of the features used in our classifier might also be biased by the prior existence of approved drugs within their molecular pathways, such as the use of PPI information^52^. Yet, the derived predicted probabilities for the extensive list of gene-trait pairs with robust evidence of a causal role now provide a nuanced and highly granular means of comparison and constitute a helpful resource to identifying the most promising candidates for downstream exploration.

## Data availability

Full summary statistics of all the two-sample MR are publicly available and can be downloaded at the VA’s Centralized Interactive Phenomics Resource (CIPHER) web portal (https://phenomics.va.ornl.gov/). All supporting annotation files are also available for download at CIPHER.

## Code availability

Code used in this manuscript can be located at the following GitHub repository: https://github.com/BrianFerolito/MVP_efficacy

## Acknowledgements

This research is based on data from the Million Veteran Program, Office of Research and Development, Veterans Health Administration, and was supported by award #MVP000. This publication does not represent the views of the Department of Veteran Affairs or the United States Government.

Full Million Veteran Program acknowledgement can be found in Supplementary File 3.

This research used resources from the Knowledge Discovery Infrastructure at the Oak Ridge National Laboratory, supported by the Office of Science of the U.S. Department of Energy under Contract No. DE-AC05-00OR22725 and the Department of Veterans Affairs Office of Information Technology Inter-Agency Agreement with the Department of Energy under IAA No. VA118-16-M-1062.

We would like to acknowledge the time and effort of the study participants and researchers in the Fenland study DOI: 10.22025/2017.10.101.00001; https://www.mrc-epid.cam.ac.uk/research/studies/fenland/.

JCW is funded by the UK Medical Research Council via programme grant MC_UU_00002/18

We would like to acknowledge Mohd Karim who aided in prior work that laid the foundation for this study.

## Contributions

B.R.F., J.P.C., A.P. conceived and designed the project.

I.A.S, B.Z., A.L., M.G., Y.T., F.H., H.I. consulted and helped to develop drug related data.

C.G., D.R., H.D. curated and assembled QTL data.

B.R.F., D.J.G., A.C.P., G.M.P., H.D., Y.L., G.P., G.G.T, G.B. developed code for various modules of pipeline.

J.W., A.H., K.C., J.M.G, T.G., G.H. provided feedback on experimental design.

A.S.B., C.L., J.C.W., E.D., L.S. provided substantive feedback on the manuscript.

S.M., J.M., J.H., S.D., S.W., K.L., T.C., P.T. provided administrative and material support.

R.M., L.C., N.K. helped plan and execute the project.

All authors read and approve the manuscript. Ethics declarations

## References

1. Gaziano, J. M. et al. Million Veteran Program: A mega-biobank to study genetic influences on health and disease. J Clin Epidemiol (2016) doi:10.1016/j.jclinepi.2015.09.016.

2. Sudlow, C. et al. UK Biobank: An Open Access Resource for Identifying the Causes of a Wide Range of Complex Diseases of Middle and Old Age. PLoS Med 12, (2015).

3. Pan-UKB team. https://pan.ukbb.broadinstitute.org. (2020).

4. Kurki, M. I. et al. FinnGen provides genetic insights from a well-phenotyped isolated population. Nature 613, (2023).

5. Zhou, W. et al. Global Biobank Meta-analysis Initiative: Powering genetic discovery across human disease. Cell Genomics 2, (2022).

6. Dhindsa, R. S. et al. Rare variant associations with plasma protein levels in the UK Biobank. Nature 622, 339–347 (2023).

7. Sun, B. B. et al. Plasma proteomic associations with genetics and health in the UK Biobank. Nature 622, 329–338 (2023).

8. Santos, R. et al. A comprehensive map of molecular drug targets. Nat Rev Drug Discov 16, (2016).

9. Pividori, M. et al. Projecting genetic associations through gene expression patterns highlights disease etiology and drug mechanisms. Nat Commun 14, 5562 (2023).

10. Duffy, Á. et al. Development of a human genetics-guided priority score for 19,365 genes and 399 drug indications. Nat Genet (2024) doi:10.1038/s41588-023-01609-2.

11. Labrecque, J. & Swanson, S. A. Understanding the Assumptions Underlying Instrumental Variable Analyses: a Brief Review of Falsification Strategies and Related Tools. Curr Epidemiol Rep 5, 214– 220 (2018).

12. Walker, V. M., Zheng, J., Gaunt, T. R. & Smith, G. D. Phenotypic Causal Inference Using Genome-Wide Association Study Data: Mendelian Randomization and Beyond. Annu Rev Biomed Data Sci 5, 1–17 (2022).

13. Burgess, S. et al. Using genetic association data to guide drug discovery and development: Review of methods and applications. Am J Hum Genet 110, 195–214 (2023).

14. Trajanoska, K. et al. From target discovery to clinical drug development with human genetics. Nature 620, 737–745 (2023).

15. Verma, A. et al. Diversity and scale: Genetic architecture of 2068 traits in the VA Million Veteran Program. Science (1979) 385, (2024).

16. Hamosh, A., Scott, A. F., Amberger, J. S., Bocchini, C. A. & McKusick, V. A. Online Mendelian Inheritance in Man (OMIM), a knowledgebase of human genes and genetic disorders. Nucleic Acids Res 33, (2005).

17. Karczewski, K. J. et al. Systematic single-variant and gene-based association testing of thousands of phenotypes in 394,841 UK Biobank exomes. Cell Genomics 2, (2022).

18. Blake, J. A. et al. Mouse Genome Database (MGD): Knowledgebase for mouse-human comparative biology. Nucleic Acids Res 49, (2021).

19. Zdrazil, B. et al. The ChEMBL Database in 2023: a drug discovery platform spanning multiple bioactivity data types and time periods. Nucleic Acids Res 52, D1180–D1192 (2024).

20. Zhao, J. H. et al. Genetics of circulating inflammatory proteins identifies drivers of immune-mediated disease risk and therapeutic targets. Nat Immunol 24, 1540–1551 (2023).

21. Keefe, D. L. Trastuzumab-associated cardiotoxicity. Cancer 95, 1592–600 (2002).

22. Nelson, M. R. et al. The support of human genetic evidence for approved drug indications. Nat Genet 47, 856–60 (2015).

23. King, E. A., Davis, J. W. & Degner, J. F. Are drug targets with genetic support twice as likely to be approved? Revised estimates of the impact of genetic support for drug mechanisms on the probability of drug approval. PLoS Genet 15, e1008489 (2019).

24. Pavlides, J. M. W. et al. Predicting gene targets from integrative analyses of summary data from GWAS and eQTL studies for 28 human complex traits. Genome Med 8, 84 (2016).

25. Sadler, M. C., Auwerx, C., Deelen, P. & Kutalik, Z. Multi-layered genetic approaches to identify approved drug targets. Cell genomics 3, 100341 (2023).

26. Yang, C. et al. Mendelian randomization and genetic colocalization infer the effects of the multi-tissue proteome on 211 complex disease-related phenotypes. Genome Med 14, 140 (2022).

27. Zuber, V. et al. Combining evidence from Mendelian randomization and colocalization: Review and comparison of approaches. Am J Hum Genet 109, 767–782 (2022).

28. Zhou, Y. et al. A comprehensive SARS-CoV-2–human protein–protein interactome reveals COVID-19 pathobiology and potential host therapeutic targets. Nat Biotechnol 41, (2023).

29. Meldal, B. H. M. et al. Complex Portal 2018: Extended content and enhanced visualization tools for macromolecular complexes. Nucleic Acids Res 47, (2019).

30. Luck, K. et al. A reference map of the human binary protein interactome. Nature 580, (2020).

31. Rolland, T. et al. A proteome-scale map of the human interactome network. Cell 159, (2014).

32. Türei, D., Korcsmáros, T. & Saez-Rodriguez, J. OmniPath: Guidelines and gateway for literature-curated signaling pathway resources. Nature Methods vol. 13 Preprint at 10.1038/nmeth.4077 (2016).

33. Alanis-Lobato, G., Andrade-Navarro, M. A. & Schaefer, M. H. HIPPIE v2.0: Enhancing meaningfulness and reliability of protein-protein interaction networks. Nucleic Acids Res 45, (2017).

34. Schaefer, M. H. et al. Hippie: Integrating protein interaction networks with experiment based quality scores. PLoS One 7, (2012).

35. Schaefer, M. H. et al. Adding Protein Context to the Human Protein-Protein Interaction Network to Reveal Meaningful Interactions. PLoS Comput Biol 9, (2013).

36. Suratanee, A. et al. Characterizing Protein Interactions Employing a Genome-Wide siRNA Cellular Phenotyping Screen. PLoS Comput Biol 10, (2014).

37. Szklarczyk, D. et al. STRING v11: Protein-protein association networks with increased coverage, supporting functional discovery in genome-wide experimental datasets. Nucleic Acids Res 47, (2019).

38. Scannell, J. W. et al. Predictive validity in drug discovery: what it is, why it matters and how to improve it. Nat Rev Drug Discov 21, 915–931 (2022).

39. Wang, Y. et al. Rare variants in long non-coding RNAs are associated with blood lipid levels in the TOPMed whole-genome sequencing study. Am J Hum Genet 110, 1704–1717 (2023).

40. Cadby, G. et al. Comprehensive genetic analysis of the human lipidome identifies loci associated with lipid homeostasis with links to coronary artery disease. Nat Commun 13, 3124 (2022).

41. Graham, S. E. et al. Author Correction: The power of genetic diversity in genome-wide association studies of lipids. Nature 618, E19–E20 (2023).

42. Seidah, N. G. et al. Annexin A2 is a natural extrahepatic inhibitor of the PCSK9-induced LDL receptor degradation. PLoS One 7, e41865 (2012).

43. Mayer, G., Poirier, S. & Seidah, N. G. Annexin A2 is a C-terminal PCSK9-binding protein that regulates endogenous low density lipoprotein receptor levels. J Biol Chem 283, 31791–801 (2008).

44. Zhu, Z. et al. Integration of summary data from GWAS and eQTL studies predicts complex trait gene targets. Nat Genet 48, 481–7 (2016).

45. Zheng, J. et al. Phenome-wide Mendelian randomization mapping the influence of the plasma proteome on complex diseases. Nat Genet 52, 1122–1131 (2020).

46. Zhao, H. et al. Proteome-wide Mendelian randomization in global biobank meta-analysis reveals multi-ancestry drug targets for common diseases. Cell genomics 2, None (2022).

47. Ferkingstad, E. et al. Large-scale integration of the plasma proteome with genetics and disease. Nat Genet 53, 1712–1721 (2021).

48. Ritchie, M. D., Holzinger, E. R., Li, R., Pendergrass, S. A. & Kim, D. Methods of integrating data to uncover genotype-phenotype interactions. Nature Reviews Genetics vol. 16 Preprint at 10.1038/nrg3868 (2015).

49. Xiong, X. et al. Knowledge-Driven Online Multimodal Automated Phenotyping System. medRxiv (2023) doi:10.1101/2023.09.29.23296239.

50. Lin, Y., Lu, K., Yu, S., Cai, T. & Zitnik, M. Multimodal learning on graphs for disease relation extraction. J Biomed Inform 143, 104415 (2023).

51. Wen, J. et al. Multimodal representation learning for predicting molecule-disease relations. Bioinformatics 39, (2023).

52. Sadler, M. C., Auwerx, C., Deelen, P. & Kutalik, Z. Multi-layered genetic approaches to identify approved drug targets. Cell genomics 3, 100341 (2023).

53. Duffy, Á. et al. Development of a human genetics-guided priority score for 19,365 genes and 399 drug indications. Nat Genet 56, 51–59 (2024).

54. Minikel, E. V., Painter, J. L., Dong, C. C. & Nelson, M. R. Refining the impact of genetic evidence on clinical success. Nature 629, (2024).

