## Supplementary material for "Leveraging Large-Scale Biobanks for Therapeutic Target Discovery": Methods

**ONLINE METHODS**

**Phenotypes**

The comprehensive Million Veteran Program^1^ (MVP) Genome-wide Association Study x Phenome-wide Association Study (gwPheWAS) produced GWAS summary statistics for a large number of phenotypes. These phenotypes largely fit into four major categories. The first of these are clinical outcomes in the electronic health records (EHR) which are represented by phecodes. Also included were lab values, vital status measurements, and questionnaire data. There were 1,893 phenotypes available for analysis (Supplementary Table 1).

**GWAS in MVP**

Genome-wide association studies were conducted in MVP participants using a mixed model approach in SAIGE^2^ adjusting for age, sex, and first 10 principal components as implemented as part of the MVP genome-wide PheWAS project. For EHR-based values, cases and controls were classified by phecodes, which comprise a high-throughput phenotyping tool based on International Classification of Diseases (ICD) codes^3^. Individuals needed at least two phecode-mapped ICD codes to be phenotyped. Briefly, quantitative traits were inverse normal transformed to satisfy the normality assumptions before adjusting for covariates and outliers falling six standard deviations from the mean were excluded. The lab data come from clinician-adjudicated clinical labs with all values falling within specific ranges. For each lab type, the minimum, mean, and maximum values were taken with both raw values and inverse normalized values used. Results were filtered to remove variants with poor imputation quality (R2<0.3) or that were very rare (minor allele frequency (MAF) < 0.01%) or minor allele count (MAC) > 20. Race and ethnicity were stratified using HARE^4^, a supervised learning algorithm which uses both self-identified race and ethnicity as well as genetically inferred ancestry. Detailed methods can be found at [Diversity and scale: Genetic architecture of 2068 traits in the VA Million Veteran Program | Science](https://www.science.org/doi/10.1126/science.adj1182)^5^.

**Harmonization of MVP, UK Biobank, and FinnGen for meta-analysis**

To harmonize phenotypes between MVP, the Pan-UK Biobank (UKBB)^6^, and FinnGen^7^ (version 10), disease-based traits were mapped using codes provided by the biobanks. MVP used entirely phecodes (1,171), while the UK Biobank used phecodes (1,327) as well as ICD10 codes (915) to label their disease phenotypes. After restricting studies in the UK Biobank to those with European ancestry in their list of populations, all possible direct phecode to phecode (1,013) matches were made between the biobanks (Supplementary Table 1). The sample size for each phenotype from each biobank is also available in Supplementary Table 1. UKBB traits with an ICD10 code were then mapped to phecodes using a conversion table^8^, and if the derived phecode had not already been mapped to MVP in the previous step, then a match was made if possible (n = 10). For MVP phecodes that remained unmapped to UKBB, studies with case counts higher than 4,000 were reviewed and a decision was made on a case-by-case basis to map manually (n = 53) by a clinician before aligning with FinnGen. For all unmapped MVP phecodes with case counts lower than 4,000, MVP only data was used in for analysis. ^9^To map MVP and UK Biobank phenotypes to FinnGen, we manually mapped all available R10 FinnGen phenotypes to those previously available from MVP or UKBB. ^10^All matches were double checked through clinical adjudication and comparison statistics using the number of cases per resource for each phenotype to identify significant outliers from this mapping strategy (See Supplementary Appendix 4 on Biobank Harmonization). Situations were a significant deviation from the overall relationship between MVP/UKBB/FinnGen case counts was observed were analyzed on a one-by-one case and a final harmonization decision was made. This led FinnGen to being mapped only with MVP (32 instances), only with UKBB (174 instances), and with both MVP and UKBB (501 instances).

**Meta-analysis between MVP, UKBB and FinnGen**

Pan-UKBiobank GWAS physical positions were converted from genome build GRCh37 to GRCh38 using LiftOver^9^. Using METAL^11^, we performed fixed effects inverse-variance weighted meta-analysis of MVP European results, UKBB European results, and FinnGen and obtained estimates of heterogeneity (option in METAL: ANALYZE HETEROGENEITY). If mapping was not possible between MVP, UKBB, or FinnGen, then only MVP results or select UKBB phenotypes were retained for further analyses. For very small p-values in MVP which were set to zero, we set to the lowest possible decimal place allowed in Python using sys.float_info.min() (2.2250738585072014e-308). To test for inflation of p-values following meta-analysis, we calculated the lambda values for all meta-analysis results. Any phenotype having a lambda value greater than 1.15 (n = 289) was rerun including the genomic control parameter in METAL. Phenotypes that were not meta-analyzed were also tested for inflated p-values. We found 81 phenotypes that needed correction, the vast majority being labs (75 studies). Due to highly inflated p-values, we removed eight height-based phenotypes from consideration. Lambda values can be found in Supplementary Table 14.

**Instruments for MR:**

We have used 5 different sources of genetic instruments in the present work. Each source was analyzed independently, that is, no meta-analysis or joint analysis among sources was conducted. MR results from different sources were then analyzed using the rule-based system provided (for more information, please refer to subsection on the use of different genetic instrument sources for MR analysis). Below we describe the instrument selection criteria for each source. Finally, an overall descriptive analysis of used genetic instruments can be found in the Supplementary Appendix I.

**GTEx version 8**

Independent *cis*-eQTLs were identified per gene by performing up to 5 conditional analyses in regions (+/-1 Mb from the transcription start site [TSS] of each gene) using GTEX v8^12^ individual-level data, additionally adjusting for the peak variant if there exists an association reaching a p-value of 1e-4. The primary signal is unconditional. A total of 14,752 GTEx GENCODE version 26 genes were considered for the next steps (<https://storage.googleapis.com/gtex_analysis_v8/reference/gencode.v26.GRCh38.genes.gtf>).

To identify the independent signals, we considered primary and conditional associations passing a p-value < 5e-8. We then extracted estimates of effect size and standard errors from the unconditional association to use in the next steps of Mendelian Randomization. This approach was taken for each available GTEx tissue. We used 120,303 markers instrumenting 24,116 genes.

**eQTLGen**

Summary statistics files were downloaded from eQTLGen^13^ <https://eqtlgen.org/cis-eqtls.html>,

Since only Z-scores and p-values, but not the betas, are listed, we have computed the beta and SE from the following formula: Beta = z / sqrt(2p(1− p)(n + z^2)); SE =1 / sqrt(2p(1− p)(n + z^2)), after downloading the MAF file provided here: https://molgenis26.gcc.rug.nl/downloads/eqtlgen/cis-eqtl/2018-07-18_SNP_AF_for_AlleleB_combined_allele_counts_and_MAF_pos_added.txt.gz.

The summary statistics reported SNP-gene associations (<1Mb from the center of the gene and tested in at least 2 cohorts) across 19,250 genes (17,114 in common between GTEx and eQTLGen). We defined instruments using the smallest p-value per gene.

We used liftOver to convert SNPs from GRCh37 to GRCh38 genome build. There were 558 SNPs in GRCh37 that could not be converted and were dropped from our analysis. We used 10,669 markers instrumenting 11,175 genes.

**deCODE**

We downloaded the published GWAS of SOMAscan v4 in 35,000 individuals of European ancestry for 4,907 aptamers from deCODE^14^ (<https://www.decode.com/summarydata/>). We subset Table S02 from the downloaded document to only cis-pQTLs and removed duplicates by chromosome and position, resulting in 5,662 instruments across 1,663 genes encoding a protein (1,674 proteins and 1,703 SeqIDs). We used 4,775 markers instrumenting 1,624 genes.

**Fenland**

We obtained pQTLs directly from the Fenland study^15^ , a genome-proteome-wide association study among 10,708 participants of European-descent conducted using 10.2 million genetic variants and plasma abundances of 4,775 distinct protein targets (proteins targeted by a least one aptamer) measured using the SOMAscan V4 assay on 4,979 aptamers (4,775 unique protein targets). We used Table S2 reporting significant genetic variant pQTLs defined as passing a Bonferroni threshold of p < 1.004e-11 and secondary signals using approximate conditional analysis for each genomic region identified by distance-based clumping of association statistics. A total of 2,900 *cis*-pQTLs across 1,557 genes (mean = 1.9, min = 1, max = 14) covering an equal number of proteins from the Fenland study were used as proposed instruments. The unconditional summary statistics from this study are available in an open resource platform ([www.omicscience.org](http://www.omicscience.org)). We used 2,881 markers instrumenting 1,510 genes.

**ARIC**

We downloaded the published *cis*-pQTL GWAS from the Atherosclerosis Risk in Communities (ARIC) study^16^ (<http://nilanjanchatterjeelab.org/pwas/>), which contains SOMAscan v4 on 4,657 plasma proteins measured in 7,213 European American individuals. The ARIC study is a prospective study conducted initially from 1987 to 1989 in four communities across the U.S.: Washington County, Maryland; suburbs of Minneapolis, Minnesota; Forsyth County, North Carolina; and Jackson, Mississippi. Blood samples for plasma protein data were collected from participants in the third visit in 1993 to 1995. SOMAmers that mapped to multiple gene targets or without a position record in the BioMart database for the target protein-coding gene or without any SNPs in the *cis* region were excluded from further analysis. The *cis*-region is defined as ±500 kb of the TSS of the target protein-coding gene in the *cis*-pQTL analysis. Genotyping of ARIC samples was performed on the Affymetrix 6.0 DNA microarray and imputed to the TOPMed reference panel (Freeze 5b). SNPs with minor allele frequency (MAF) below 1%, imputation quality (R2) below 0.8, call rates less than 90%, or Hardy-Weinberg equilibrium P values below 10^-6 were removed from further analysis. A total of 2,004 significant SOMAmers were therefore identified in the original study that had at least one *cis*-pQTL (FDR < 5%) near the gene of the putative protein. We used unconditional estimates from this list of 2,004 cis-pQTLs from the original study. We used 1,612 markers instrumenting 1,594 genes.

**Mendelian Randomization**

Two-sample Mendelian Randomization (MR) of each of 16,915 protein-coding genes were performed against all phenotypes using instruments from the five sources of eQTLs and pQTLs. The datasets used for instruments provided summary statistics for the unconditional primary association. For each of the datasets described above, we used the instruments identified by the authors. We extracted the corresponding effect size and standard errors from the unconditional association to use in MR. In order to determine the correct ordering of alleles between the datasets we utilized the harmonise_data() function from the TwoSampleMR^17^ package in R^18^.

We used the Wald Ratio for instruments with one genetic variant and inverse variance weighted MR for instruments with multiple genetic variants. We additionally performed MR-Egger for proteins/expression with three or more instruments to be used as a sensitivity analysis. We tested for heterogeneity across variant-level MR estimates, using the Cochrane Q method (mr_heterogeneity option in TwoSampleMR package) and the MR-Egger intercept.

We define genes with significant MR as genes with p-value ≤ 1.6e-9 5e-8 for any MR test. This value was obtained by dividing 0.05 by the number of unique gene-trait pairs tested in our study (31,525,236 unique gene-traits). If a gene was significant in multiple QTL sources, we required the directionality of the betas to be concordant. If a gene passed in both eQTL and pQTL sources, we only considered the pQTL (more details on the next section).

**On the use of different genetic instrument sources for MR analysis**

As detailed, we have used two different sources for eQTL instruments (GTEx and eQTLGen) and three different sources for pQTL instruments (ARIC, DeCODE, and Finland). Because prior studies have shown discrepancy between pQTLs and eQTLs, and because the specific methods of derivation are not exactly the same for each study, we have devised a set of inclusion rules of the genome-wide significant MR results that were further characterized and annotated. This arbitrary set of criteria was designed with the intent to maximize the use of the available eQTL and pQTL information in our used sources, while restricting to downstream analyzes only results that were concordant among, but not necessarily between, the different source types. First, we did not meta-analyze or combine different sources to create multi-snp instrument for a gene. As such, for each gene and phenotype, MR analysis was conducted for each source (for GTEx we conducted this analysis within each represented tissue). As a result, multiple MR estimates are available for each tested gene-trait. To select a particular gene-trait for downstream analysis, we only selected significant gene-traits with concordant results in regards to the predicted directionality of effect from MR. To be more specific, if a gene-trait was only seen as significant because of one simple instrument source, it was accepted. If MR was significant in more than one source we applied the following rules: (1) for gene pairs that were significant for both at least one eQTL and at least one pQTL sources, we only check concordance of directionality among pQTL sources (acknowledging that eQTL and pQTL genetic proxies might be discordant due to the effects of post-transcriptional regulation); (2) the signal of the MR beta estimate had to be concordant among all significant MR results for the tested gene pair (including all different tissues from GTEx in case more than one was significant); (3) all discordant gene-traits were excluded from further consideration.

It is, thus, important to consider that all derived causal effect estimates are assumed to be associated with the specific molecular mediator being instrumented. That is, the transcriptional level of a gene or the expression level of a protein in whole blood, depending on the used instrument. However, there are well described situations where gene expression is regulated by the same genetic variant, but differently, and in opposite directions, at different tissues; or when RNA transcript and protein expression are regulated by the same genetic variant, but in opposite directions. Conservatively, we required concordant results from all significant MR results from all different GTEx tissues, reducing the possibility of misspecification due to the former described scenario. However, by considering only pQTL results when these were available we have not excluded gene-traits that could have opposite results at the eQTL and pQTL levels. A direct comparison of the correlations between each pairwise combination of significant MR beta results can be seen on Supplementary Figure 6. The number of selected and excluded gene-traits on each of the steps of the selection process is illustrated on Supplementary Figure 1.

**Colocalization**

For genes with a significant MR result we performed colocalization between the outcome GWAS (or meta-analysis) and the *cis*-variants available from the QTL sources passing MR for that gene-trait using the coloc package^19^ in R. Marginal (unadjusted) e/pQTL results and unconditional results on each of the instruments used in the MR were used. For GTEx instruments identified by conditional associations, since the full associations were available to us, the conditional results were used. We used variants with MAF > 1% and a +/-250KB window around each of the instruments. We defined strong colocalization as having posterior probability for hypothesis 4 (PP.H4) > 0.8 (the probability of a shared causal variant) for at least one instrumental variant. We also noted when significant gene-trait pairs had strong colocalization for every instrumental variant.

For case-control studies we incorporated p-values and the proportion of the samples that are cases in the outcome GWAS. For quantitative traits, sdY (standard deviation of the trait) was calculated using the variance of beta and MAF. Prior probabilities were all set to default values.

**Assessment of Druggability**

We extracted drug information for protein targets from ChEMBL^20^ (v34). For all protein targets, we acquired Ensembl IDs when available using UniProt’s rest API. For each drug where the information was available, we assigned all indications for those drugs and the clinical phase for that indication. Also added was the mechanism of action (MoA) for the drug and the interaction it has with the target. For this we classified as positive modulation (activator) or negative modulation (inhibitor), or other. Drug information can be found in Supplementary Table 15.

**Protein-protein Interactions**

Using the approach introduced by MacNamara et al^21^, we used protein-protein interactions (PPI) to investigate associations between significant gene-traits and approved drug target-indications. We constructed a PPI network containing 1-step interactions between all instrumented genes. The aggregated protein-protein interactions network was constructed containing seven different PPI resources: Complex^22^, Lit BM^23,24^, Metabase (https://portal.genego.com/), OmniPath^25^, HIPPIE^26–29^, HI union^24^, and STRING^30^. In this process, we used the .8 quantile of the STRING scores as the significance cutoff and only incorporated the interactions with scores within the top 20 percentile. A similar significance cutoff was calculated and applied for the HIPPIE dataset.

**Calculation of pairwise semantic distance between terms**

Here we describe the method and implementation for the calculation of pairwise semantic distance between trait terms. The dataset consists of trait terms from the eight following source groups: Clinvar, Drug indications (ChEMBL34), GWAS Catalog, Knockout models (MGI database), genetic phenotypes from MVP, UKB, and FinnGen, OMIM, and pLOF burden analysis from UKBB. We used ScispaCy^31^ (en_core_sci_lg version 0.5) as the encoder model for its ability to efficiently encode biomedical text and symbols, and for each of the terms we computed a 200 x 1 vector as the semantic vector representation of the term label. The generated encodings were then loaded into an Elasticsearch vector store as dense vectors, where the top-N candidates that are most closely associated with the input vector of a query term can be efficiently computed and retrieved on-the-fly using a k-Nearest Neighbour (kNN) search based on a cosine similarity metric. For each of these trait terms, we then computed the cosine similarities between this trait term and each term of the eight source groups (including the group this trait term belongs to). We then retrieved, for each trait term their 3% most similar trait terms in the semantic vector space for each source. Source code for the implementation is available under the "MRCIEU/phenotype-mapping" repository on GitHub here https://github.com/MRCIEU/phenotype-mapping/tree/2023-06-mvp-terms/analysis/pipelines/mvp_ontology_distance_round3.

**Novelty allocation using GWAS Catalog**

To assess one novelty aspect of our results we consulted the GWAS Catalog^32^ (downloaded with most recent update from 1/30/2023) to determine if the gene-trait pair found to be significantly associated in our MR results had a gene/variant previously reported as associated with the phenotype in question. To map our phenotypes (phecodes) to the ontology-based phenotypes, we used an NLP tool to help rank and assign matches between the description of the phenotype and EFO ontology terms. The full list of EFO terms used and their parent terms is found in Supplementary Table 16. We also assigned parent terms, or high-level disease classification by using the assigned EFO terms and ascending ancestor terms until we captured a predefined ontology term in our list (Supplementary Table 13). Additionally, for each phenotype we created a list of top 3% closest phenotypes in the GWAS catalog by the embedding method described previously.

For gene-trait pairs that passed two-sample MR we selected the region corresponding to 250KB before the TSS of the gene in question to 250KB after the TSE of the gene in question. We then searched the table for either a direct match on ontology, a match on the parent term, or by a semantic distance match. For each type of match (self, parent, and distance) we provide a score, 1 for a match on the region, and 0 for no match and therefore novel. It should be highlighted that this search scheme is not error proof. GWAS catalog significant associations sometimes do not reflect the full body of literature linking genetic variants in a gene to a specific phenotype.

**Evaluating features associated with selected MR findings**

To evaluate whether specific features of our data could be predictive of a selected MR gene-trait, we have created a file containing all selected gene-traits by MR (after described filter steps) and the same number of non-selected MR results. All gene-traits (selected MR and non-selected MR gene-traits) were mapped to features thought to be predictive of a significant MR result (please refer to Supplementary Table 11for a description of each engineered feature). Matching of significant and non-significant results followed the same filtering steps as used for significant results, including checks on directionality and eQTL and pQTL sources. Specifically, for each of the final selected MR results, we randomly selected a result without robust evidence of causality with the same number of eQTL and/or pQTL sources to test for directionality of the estimated effect. The specific QTL source(s) selected to be represented in the non-selected MR result was a function of the number of sources represented in the positive result and a weighted probability reflecting the relative distribution of each QTL source in the overall tested MR gene-traits. The resultant file was then used to compare feature enrichment among significant MR results. Feature enrichment was always estimated using a logistic regression approach in which the outcome was being a significant gene-trait in the MR analysis or not. Enrichment of each derived feature was tested using the above described set of gene-traits in a logistic regression framework using as dependent variable the status of the MR gene-trait (initially selected or non-selected) and as the independent variable each derived feature.

**Mapping Orthogonal Sources**

To garner support for our significant gene-trait pairs, we sought to identify whether these connections had been previously uncovered in an assortment of sources of biological information. To determine if our gene-trait pair had been reported at both the phenotype and gene level, we observed OMIM^33^, putative Loss of Function data from GeneBass^34^, knockout models from MGI^35^, and variants of clinical significance from ClinVar^33,36^. For each source, we assigned EFO codes for the phenotypes as well as the parent terms for the selected phenotypes. The OnToma package for Python was used to map OMIM and pLOF phenotypes to EFO codes and terms, MONDO codes and terms, and ORDO/Orphanet codes and terms. We have referred to any ontology terms from these systems as EFO codes. The OnToma package (<https://github.com/opentargets/OnToma> v1.1.0) supports two forms of input: phenotype descriptions such as “heart failure” and phenotype codes from non-EFO systems, such as OMIM IDs. The method then returns a list of EFO codes matching the input if any are found. OnToma mapping was also attempted with phenotypes from MGI, but only a small subset of phenotypes mapped successfully. Therefore, MGI phenotypes were mapped to EFO terms at the distance and parent level only.

**Assessment of instruments: PVE and F-statistic calculations**

We compute two key parameters from the first-stage regression of the exposure phenotype on the genetic variant: the proportion of variance explained (PVE) and the F-statistic. These parameters are indicative of the power and strength of the instrumental variables (IVs) used in the study.

. The PVE^37^ by a given SNP can be expressed as a function of the effect size estimate $\left( \beta\right)$, minor allele frequency $\left( MAF \right)$, and standard error of effect size $\left( se\left( \beta\right) \right)$ for the genetic variant, and the sample size $\left( n \right)$:

$$PVE=\frac{2\beta^{2}MAF\left( 1-MAF \right)}{2\beta^{2}MAF\left( 1-MAF \right)+\left( se\left( \beta\right) \right)^{2}2nMAF\left( 1-MAF \right)}$$

To capture the ‘strength’ of the IV or set of IVs, we computed the F-statistic from the first-stage regression of the exposure phenotype on the genetic variant^38^.

The F-statistic can be expressed as a function of the $PVE$, the sample size $\left( n \right)$, and the number of IVs $\left( k \right)$:

$$F=\frac{PVE\left( n-1-k \right)}{\left( 1-PVE \right)k}$$

We used a threshold of F < 10 to define a ‘weak IV’^39^. These parameters test the validity of the first IV assumption of MR, known as the relevance assumption, which states that the genetic variant is directly associated with the exposure^40^. Weak IVs can bias the effect estimates derived from MR in the presence of confounding factors that may affect the exposure-outcome relationship. Results from this annotation are available in Supplementary Table 17.

**Creating a database of approved drug targets and clinical indications for enrichment analysis of predictive features**

The creation of a framework in which the enrichment of significant MR gene-traits, as well as other biological features, can be estimated and tested under a statistical framework was accomplished following several steps. Initially, we mapped all approved (phase 4) drug targets (genes) to unique clinical indications. This step was done manually, where all drug target and indication pairs from ChEMBL34 were clustered with similar pairs. At the end of this step, we were able to define 3,565 unique drug target-indications in ChEMBL 34. Second, we mapped all approved indications to the phenotypes with available genetic association summary statistics. These steps specifically avoided that rediscoveries or repurposing opportunities were counted multiple times (for example, discovering PCSK9 association with dyslipidemia, with mean LDL levels, and with use of anti-lipidemic medications should count as a single rediscovery and not 3 distinct ones). Finally, we created a file containing positive and negative controls for training a model for approved drug indications. Positive controls are the approved drug targets-indications just described. Negative controls were paired to positive controls by the clinical indication (to avoid bias due to increased GWAS statistical power for commonly approved indications), and derived by randomly selecting a gene from the gene set with at least one significant genome-wide MR results (to avoid the bias of creating negative gene-trait pairs using genes with weak genetic instruments). For each existing positive-control we created 10 random negative-controls. Our training dataset was composed of 3,565 positive controls and 35,614 negative controls.

**Model training and testing**

In the conducted study, a supervised classification task was undertaken employing the XGBoost algorithm^41^, facilitated through the ‘xgboost’ package in R. The approach adopted involved defining a comprehensive parameter grid to facilitate the meticulous tuning of the model, aiming to optimize its predictive performance. The parameter grid encompassed a variety of hyperparameters, including the learning rate (0.01, 0.1, 0.3), maximum depth of a tree (3,6,9), minimum child weight (1), and the subsampling rate (1). Following the establishment of the parameter grid, the ‘xgb.tree’ method was employed to implement the XGBoost model (objective function binary:logistic). A grid search cross-validation technique was utilized to systematically explore the hyperparameter space, ensuring that each unique combination of hyperparameter values was evaluated to ascertain the optimal model configuration. The model was designed to minimize the error (evaluating metric) in classification that a given gene-trait is an approved drug-target and its indication. Each model’s performance was evaluated using 5-fold cross-validation steps using 80% of our positive and negative controls from the benchmark drug-indication file to train the model. A held-out sample of 20% of positive and negative controls was used in model testing and to establish the final model´s performance. This file was derived using ten negative controls for each positive control.

The model was tested and trained using the imbalanced dataset.

**Lipids Vignette**

Initially, we filtered the ChEMBL34 clinical indications to all terms pertaining dyslipidemia treatment. For this we selected all indications matching the following EFO terms: “Abnormal circulating lipid concentration”, “Combined hyperlipidemia”, “Disorder of lipid metabolism”, “familial hypercholesterolemia”, “Hypercholesterolemia”, “hyperlipidemia”, “hyperlipoproteinemia”, “Hyperlipoproteinemia type 1”, “hyperlipoproteinemia type 3”, “Hyperlipoproteinemia type 4”. Among these terms we observed 251 different drug indications with a clear molecular target mapped to an existing drug. For the recovery of lipids genes in our set of selected MR results we used the following phenotypes: Apolipo_A,, Apolipo_B, CircCholMed, Disorders of lipoid metabolism, HDLC_Max, HDLC_Max_INT, HDLC_Mean, HDLC_Mean_INT, HDLC_Min, HDLC_Min_INT, Hypercholesterolemia, Hyperlipidemia, LDLC_Max, LDLC_Max_INT, LDLC_Mean, LDLC_Mean_INT, LDLC_Min, LDLC_Min_INT, Mixed hyperlipidemia, TotChol_Max, TotChol_Max_INT, TotChol_Mean, TotChol_Mean_INT, TotChol_Min, TotChol_Min_INT. Using these traits we observed 5746 significant MR results.

Previous GWAS hits were obtained from Graham SE et al.^42^, supplementary tables 3, 5, and 18. Previous GWAS were derived from merging all unique genes annotated in supplementary tables 3, 5, and 18 (2507 genes) and then parsing this list to contain only protein coding genes (1236 genes). Protein coding genes were obtained from [www.ensembl.org](http://www.ensembl.org) on 11/10/2023. Genes with rare variants significantly associated with lipid traits were extracted from Selvaraj MS et al.^43^ Supplementary Table 4.

**References**

1.	Gaziano, J. M. *et al.* Million Veteran Program: A mega-biobank to study genetic influences on health and disease. *J Clin Epidemiol* (2016) doi:10.1016/j.jclinepi.2015.09.016.

2.	Zhou, W. *et al.* Efficiently controlling for case-control imbalance and sample relatedness in large-scale genetic association studies. *Nat Genet* **50**, 1335–1341 (2018).

3.	Bastarache, L. Using Phecodes for Research with the Electronic Health Record: From PheWAS to PheRS. *Annu Rev Biomed Data Sci* **4**, 1–19 (2021).

4.	Fang, H. *et al.* Harmonizing Genetic Ancestry and Self-identified Race/Ethnicity in Genome-wide Association Studies. *Am J Hum Genet* **105**, 763–772 (2019).

5.	Verma, A. *et al.* Diversity and scale: Genetic architecture of 2068 traits in the VA Million Veteran Program. *Science (1979)* **385**, (2024).

6.	Sudlow, C. *et al.* UK Biobank: An Open Access Resource for Identifying the Causes of a Wide Range of Complex Diseases of Middle and Old Age. *PLoS Med* **12**, (2015).

7.	Kurki, M. I. *et al.* FinnGen provides genetic insights from a well-phenotyped isolated population. *Nature* **613**, (2023).

8.	Denny, J. C. *et al.* Systematic comparison of phenome-wide association study of electronic medical record data and genome-wide association study data. *Nat Biotechnol* **31**, (2013).

9.	Kent, W. J. *et al.* The Human Genome Browser at UCSC. *Genome Res* **12**, (2002).

10.	Malone, J. *et al.* Modeling sample variables with an Experimental Factor Ontology. *Bioinformatics* (2010) doi:10.1093/bioinformatics/btq099.

11.	Willer, C. J., Li, Y. & Abecasis, G. R. METAL: Fast and efficient meta-analysis of genomewide association scans. *Bioinformatics* **26**, (2010).

12.	Aguet, F. *et al.* The GTEx Consortium atlas of genetic regulatory effects across human tissues. *Science (1979)* **369**, (2020).

13.	Võsa, U. *et al.* Large-scale cis- and trans-eQTL analyses identify thousands of genetic loci and polygenic scores that regulate blood gene expression. *Nat Genet* **53**, (2021).

14.	Ferkingstad, E. *et al.* Large-scale integration of the plasma proteome with genetics and disease. *Nat Genet* **53**, (2021).

15.	Pietzner, M. *et al.* Synergistic insights into human health from aptamer- and antibody-based proteomic profiling. *Nat Commun* **12**, (2021).

16.	Zhang, J. *et al.* Plasma proteome analyses in individuals of European and African ancestry identify cis-pQTLs and models for proteome-wide association studies. *Nat Genet* **54**, (2022).

17.	Hemani, G. *et al.* The MR-base platform supports systematic causal inference across the human phenome. *Elife* **7**, (2018).

18.	Team, R. C. R: A Language and Environment for Statistical Computing. *R Foundation for Statistical Computing* Preprint at (2021).

19.	Giambartolomei, C. *et al.* Bayesian Test for Colocalisation between Pairs of Genetic Association Studies Using Summary Statistics. *PLoS Genet* **10**, (2014).

20.	Ochoa, D. *et al.* The next-generation Open Targets Platform: reimagined, redesigned, rebuilt. *Nucleic Acids Res* **51**, (2023).

21.	MacNamara, A. *et al.* Network and pathway expansion of genetic disease associations identifies successful drug targets. *Sci Rep* **10**, (2020).

22.	Meldal, B. H. M. *et al.* Complex Portal 2018: Extended content and enhanced visualization tools for macromolecular complexes. *Nucleic Acids Res* **47**, (2019).

23.	Rolland, T. *et al.* A proteome-scale map of the human interactome network. *Cell* **159**, (2014).

24.	Luck, K. *et al.* A reference map of the human binary protein interactome. *Nature* **580**, (2020).

25.	Türei, D., Korcsmáros, T. & Saez-Rodriguez, J. OmniPath: Guidelines and gateway for literature-curated signaling pathway resources. *Nature Methods* vol. 13 Preprint at https://doi.org/10.1038/nmeth.4077 (2016).

26.	Alanis-Lobato, G., Andrade-Navarro, M. A. & Schaefer, M. H. HIPPIE v2.0: Enhancing meaningfulness and reliability of protein-protein interaction networks. *Nucleic Acids Res* **45**, (2017).

27.	Schaefer, M. H. *et al.* Hippie: Integrating protein interaction networks with experiment based quality scores. *PLoS One* **7**, (2012).

28.	Schaefer, M. H. *et al.* Adding Protein Context to the Human Protein-Protein Interaction Network to Reveal Meaningful Interactions. *PLoS Comput Biol* **9**, (2013).

29.	Suratanee, A. *et al.* Characterizing Protein Interactions Employing a Genome-Wide siRNA Cellular Phenotyping Screen. *PLoS Comput Biol* **10**, (2014).

30.	Szklarczyk, D. *et al.* STRING v11: Protein-protein association networks with increased coverage, supporting functional discovery in genome-wide experimental datasets. *Nucleic Acids Res* **47**, (2019).

31.	Neumann, M., King, D., Beltagy, I. & Ammar, W. ScispaCy: Fast and robust models for biomedical natural language processing. in *BioNLP 2019 - SIGBioMed Workshop on Biomedical Natural Language Processing, Proceedings of the 18th BioNLP Workshop and Shared Task* (2019). doi:10.18653/v1/w19-5034.

32.	Buniello A, MacArthur JAL, Cerezo M, Harris LW, Hayhurst J, Malangone C, McMahon A, Morales J, Mountjoy E, Sollis E, Suveges D, Vrousgou O, Whetzel PL, Amode R, Guillen JA, Riat HS, Trevanion SJ, Hall P, Junkins H, Flicek P, Burdett T, Hindorff LA, C. F. & H, P. The NHGRI-EBI GWAS Catalog of published genome-wide association studies, targeted arrays and summary statistics. *Nucleic Acids Res* **47**, (2019).

33.	Hamosh, A., Scott, A. F., Amberger, J. S., Bocchini, C. A. & McKusick, V. A. Online Mendelian Inheritance in Man (OMIM), a knowledgebase of human genes and genetic disorders. *Nucleic Acids Res* **33**, (2005).

34.	Karczewski, K. J. *et al.* Systematic single-variant and gene-based association testing of thousands of phenotypes in 394,841 UK Biobank exomes. *Cell Genomics* **2**, (2022).

35.	Blake, J. A. *et al.* Mouse Genome Database (MGD): Knowledgebase for mouse-human comparative biology. *Nucleic Acids Res* **49**, (2021).

36.	Landrum, M. J. *et al.* ClinVar: Improving access to variant interpretations and supporting evidence. *Nucleic Acids Res* **46**, (2018).

37.	Shim, H. *et al.* A multivariate genome-wide association analysis of 10 LDL subfractions, and their response to statin treatment, in 1868 Caucasians. *PLoS One* **10**, (2015).

38.	Brion, M. J. A., Shakhbazov, K. & Visscher, P. M. Calculating statistical power in Mendelian randomization studies. *Int J Epidemiol* **42**, (2013).

39.	Staiger, D. & Stock, J. H. Instrumental Variables Regression with Weak Instruments. *Econometrica* **65**, (1997).

40.	Davies, N. M., Holmes, M. V. & Davey Smith, G. Reading Mendelian randomisation studies: A guide, glossary, and checklist for clinicians. *BMJ (Online)* **362**, (2018).

41.	Chen, T. & Guestrin, C. XGBoost: A scalable tree boosting system. in *Proceedings of the ACM SIGKDD International Conference on Knowledge Discovery and Data Mining* vols 13-17-August-2016 (2016).

42.	Graham, S. E. *et al.* The power of genetic diversity in genome-wide association studies of lipids. *Nature* **600**, (2021).

43.	Selvaraj, M. S. *et al.* Whole genome sequence analysis of blood lipid levels in >66,000 individuals. *Nat Commun* **13**, (2022).
