## Supplementary Figure for "Leveraging Large-Scale Biobanks for Therapeutic Target Discovery"

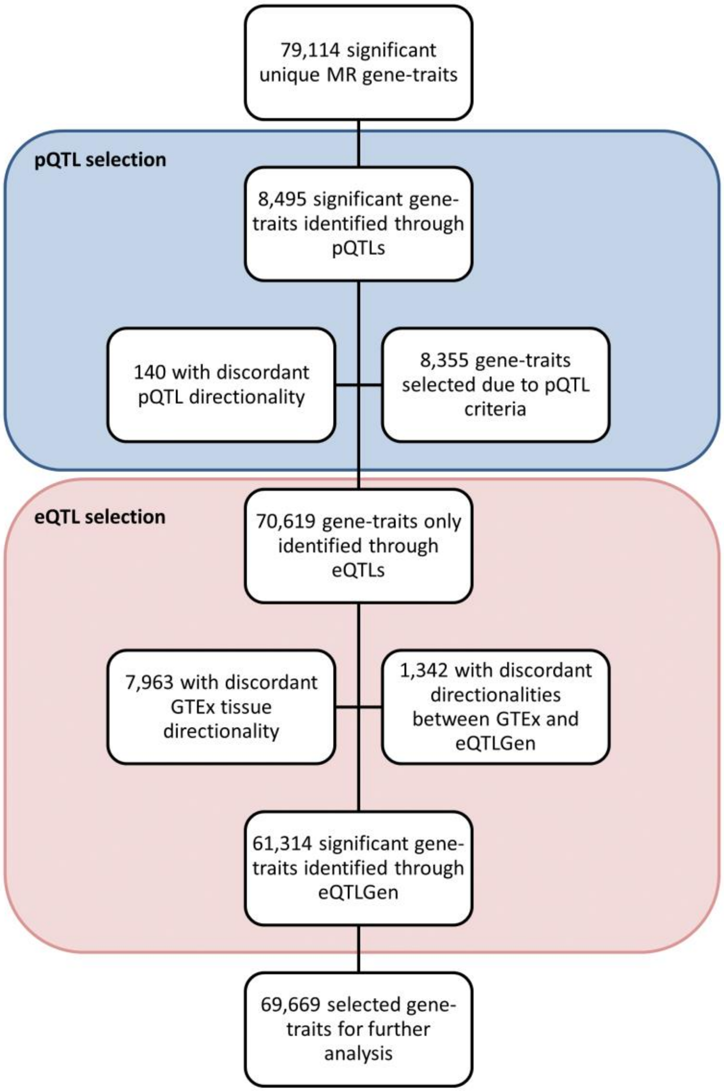


Supplementary Figure 1: Flow chart showing numbers of significant MR gene-trait pairs identified through pQTLs versus eQTLs, with additional detail pertaining to discordant and concordant directionalities.

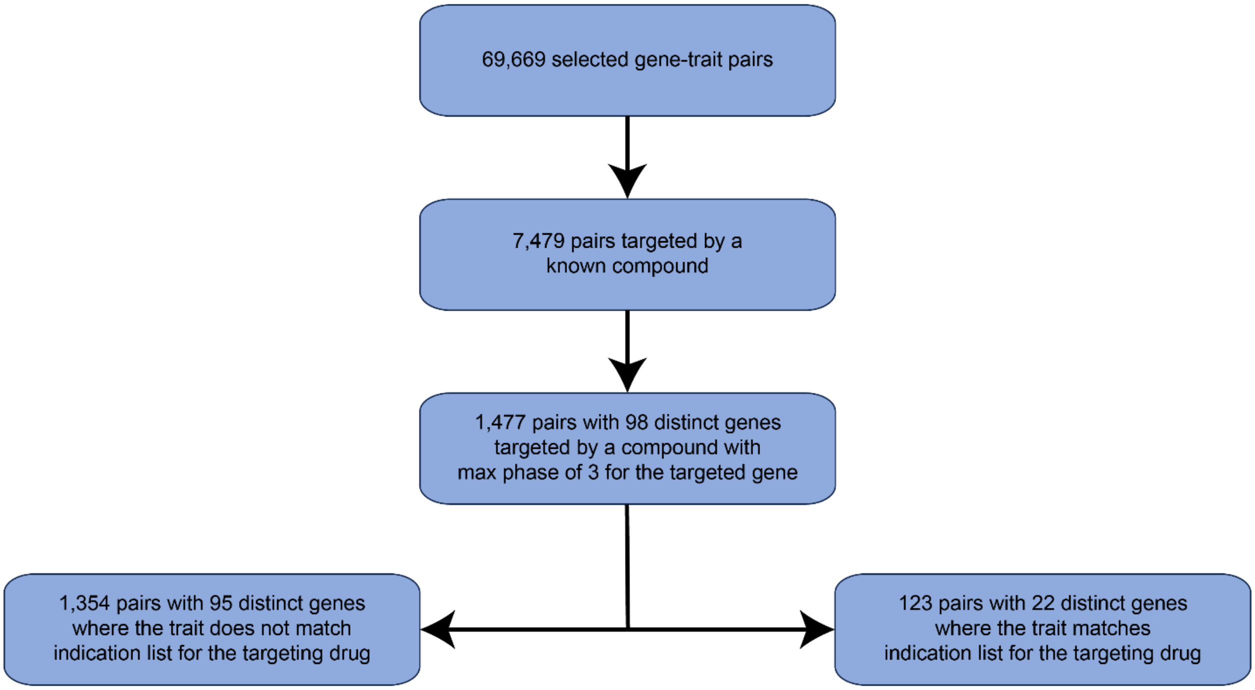


Supplementary Figure 2: Flow chart showing numbers of selected gene-trait pairs that represent rediscoveries and potential repurposing opportunities.

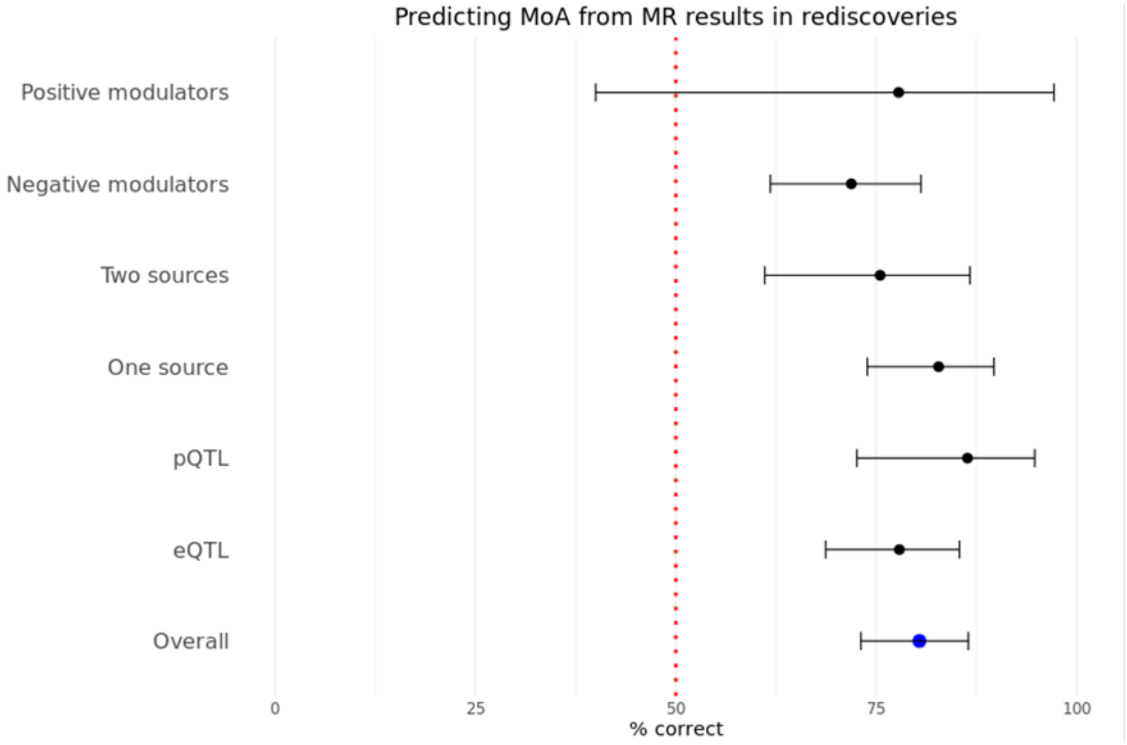


Supplementary Figure 3: Prediction of correct mechanism of action (MoA) for gene-traits identified through Mendelian Randomization (MR) for approved drugs using different filters for selecting significant MR results. Positive modulators are selected MR results only targeting drugs that are positive modulators of the target; negative modulators used selected MR results only targeting drugs that are negative modulators of the target; two sources are the selected MR results identified by more than one instrument source; one source are selected MR results only identified by a single instrument source; pQTL are selected MR results identified through the use of pQTL sources; eQTL are selected MR results identified through the use of eQTL sources.

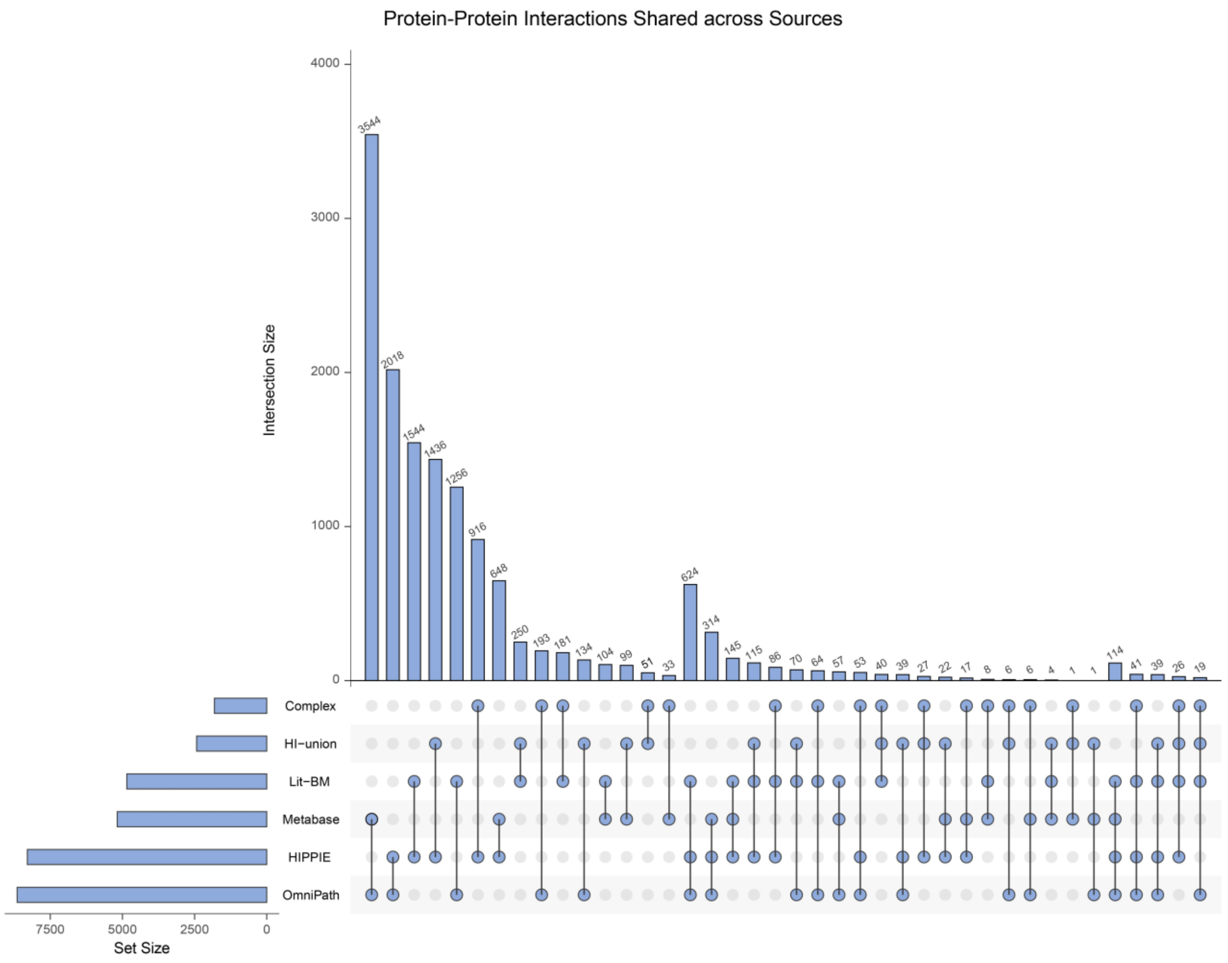


Supplementary Figure 4: Upset plot depicting the intersection between the number of protein-protein pairs among the different used protein-protein interaction (PPI) datasets.

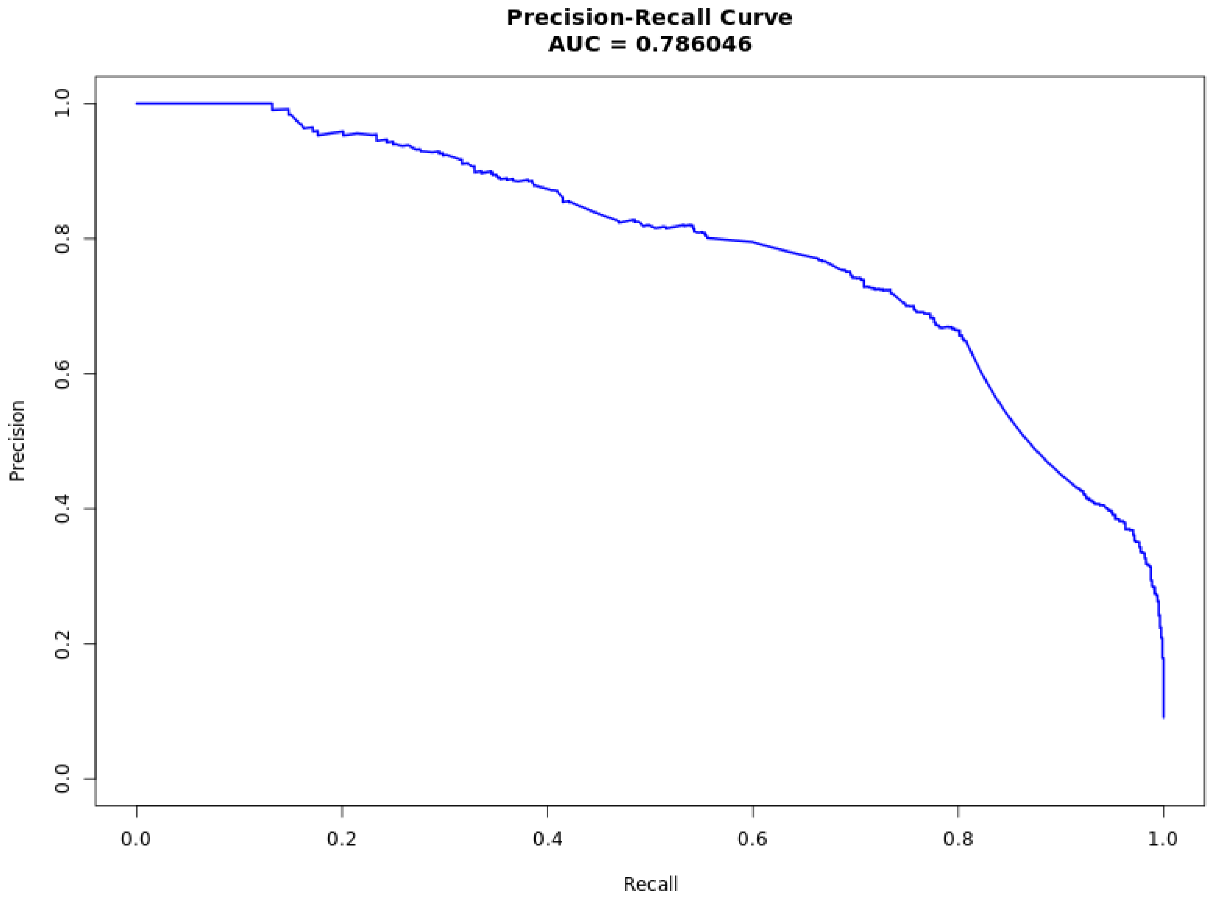


Supplementary Figure 5: Precision-recall curve for our model.

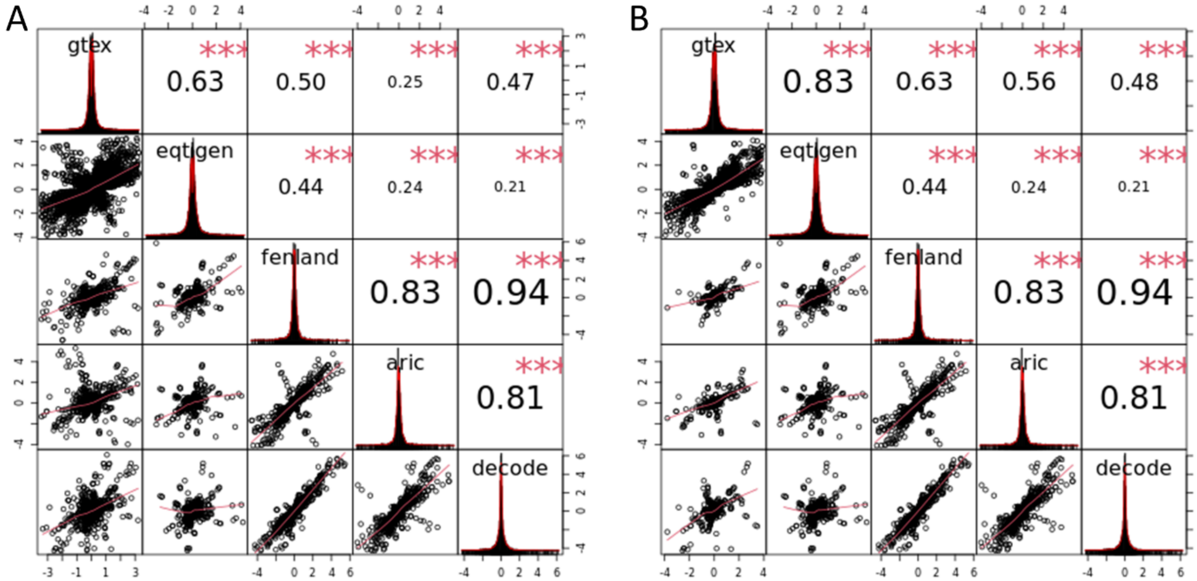


Supplementary Figure 6: Correlation matrices among the estimated MR coefficients for statistically significant traits, with panel A including all data from GTEx tissues and panel B including data only from GTEx whole blood.

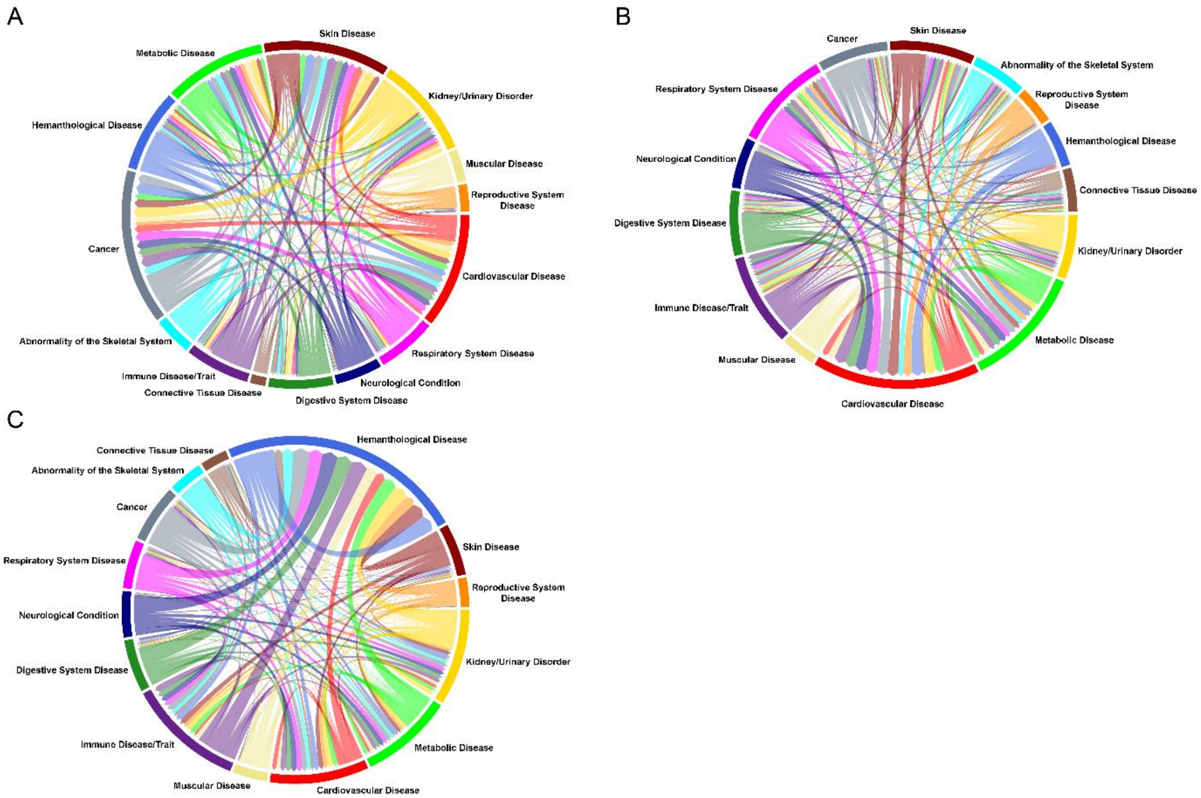


Supplementary Figure 7: Chord diagrams representing instances in which significant MR gene-traits mapped to approved drugs by parent terms - excluding lab measurement traits - were found to: panel A, increase the risk of developing deleterious traits of other parent terms (suggesting a potential safety concern); panel B, decrease the risk of developing deleterious traits of other parent terms, suggesting possible drug repurposing opportunities. In panel C, a chord diagram representing all significant MR results that mapped to an approved drug, including laboratory results. Representations for all chord diagrams are from approved drug indication parent term to MR suggested drug indication/safety concern parent term.

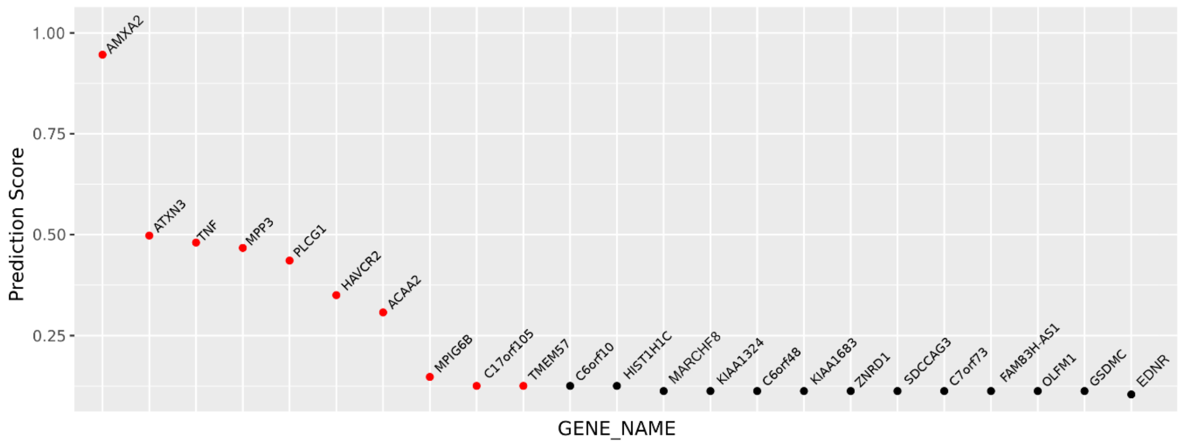


Supplementary Figure 8: Top ranked genes among all genes in a significant lipid gene-trait. Predictions were calculated using the trained classifier. Red dots are gene-trait predictions with more than 70% of a positive predictive value.

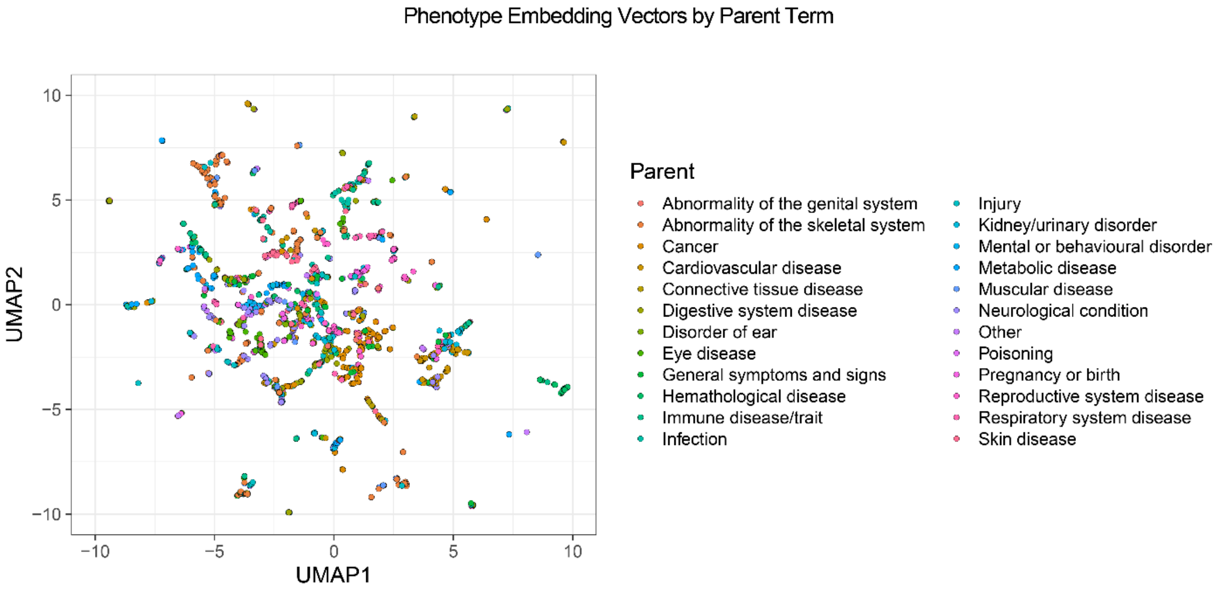


Supplementary Figure 9. UMAP representation of the distance between embedding vectors used to represent the semantic status of each studied genetic phenotype. Color representation reflects assigned parent terms.

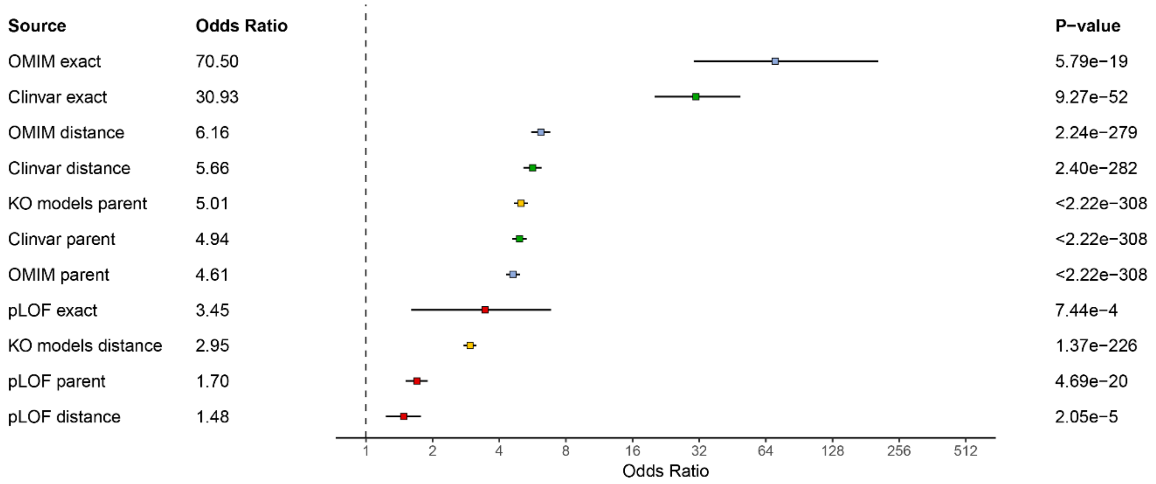


Supplementary Figure 10: Forest plot of the association between the different biological features as predictors of rediscovering a relationship with an approved drug.
